# The contribution of hospital-acquired infections to the COVID-19 epidemic in England in the first half of 2020

**DOI:** 10.1101/2021.09.02.21262480

**Authors:** Gwenan M. Knight, Thi Mui Pham, James Stimson, Sebastian Funk, Yalda Jafari, Diane Pople, Stephanie Evans, Mo Yin, Colin S. Brown, Alex Bhattacharya, Russell Hope, Malcolm G. Semple, ISARIC4C Investigators, CMMID COVID-19 working group, Jonathan M Read, Ben S Cooper, Julie V. Robotham

**Affiliations:** Centre for mathematical modelling of infectious diseases, IDE, EPH, London School of Hygiene & Tropical Medicine, London, UK; Julius Center for Health Sciences and Primary Care, University Medical Center Utrecht, Utrecht University, Utrecht, The Netherlands; Nuffield Department of Medicine, Centre for Tropical Medicine and Global Health, University of Oxford, Oxford, UK; Healthcare Associated Infections and Antimicrobial Resistance Division, National Infection Service, PHE, Colindale, London, UK; National University of Singapore Department of Medicine, Singapore; NIHR Health Protection Research Unit in Emerging and Zoonotic Infections, Institute of Infection, Veterinary and Ecological Sciences, Faculty of Health and Life Sciences, University of Liverpool, Liverpool, UK; Respiratory Medicine, Alder Hey Children’s NHS Foundation Trust, Liverpool, UK; Lancaster Medical School, Lancaster University, Lancaster, UK; NIHR Health Protection Research Unit in Healthcare Associated Infections and Antimicrobial Resistance at University of Oxford in partnership with Public Health England, Oxford, UK

**Keywords:** COVID-19, SARS-CoV-2, nosocomial transmission, mathematical modelling

## Abstract

**Background:** SARS-CoV-2 spreads in hospitals, but the contribution of these settings to the overall COVID-19 burden at a national level is unknown.

**Methods:** We used comprehensive national English datasets and simulation modelling to determine the total burden (identified and unidentified) of symptomatic hospital-acquired infections. Those unidentified would either be 1) discharged before symptom onset (“missed”), or 2) have symptom onset 7 days or fewer from admission (“misclassified”). We estimated the contribution of “misclassified” cases and transmission from “missed” symptomatic infections to the English epidemic before 31st July 2020.

**Findings:** In our dataset of hospitalised COVID-19 patients in acute English Trusts with a recorded symptom onset date (n = 65,028), 7% were classified as hospital-acquired (with symptom onset 8 or more days after admission and before discharge). We estimated that only 30% (range across weeks and 200 simulations: 20-41%) of symptomatic hospital-acquired infections would be identified. Misclassified cases and onward transmission from missed infections could account for 15% (mean, 95% range over 200 simulations: 14·1%-15·8%) of cases currently classified as community-acquired COVID-19.

From this, we estimated that 26,600 (25,900 to 27,700) individuals acquired a symptomatic SARS-CoV-2 infection in an acute Trust in England before 31st July 2020, resulting in 15,900 (15,200-16,400) or 20.1% (19.2%-20.7%) of all identified hospitalised COVID-19 cases.

**Conclusions:** Transmission of SARS-CoV-2 to hospitalised patients likely caused approximately a fifth of identified cases of hospitalised COVID-19 in the “first wave”, but fewer than 1% of all SARS-CoV-2 infections in England. Using symptom onset as a detection method for hospital-acquired SARS-CoV-2 likely misses a substantial proportion (>60%) of hospital-acquired infections.

**Funding:** National Institute for Health Research, UK Medical Research Council, Society for Laboratory Automation and Screening, UKRI, Wellcome Trust, Singapore National Medical Research Council.

**Research in context:** *Evidence before this study:* We searched PubMed with the terms “((national OR country) AND (contribution OR burden OR estimates) AND (“hospital-acquired” OR “hospital-associated” OR “nosocomial”)) AND Covid-19” for articles published in English up to July 1st 2021. This identified 42 studies, with no studies that had aimed to produce comprehensive national estimates of the contribution of hospital settings to the COVID-19 pandemic. Most studies focused on estimating seroprevalence or levels of infection in healthcare workers only, which were not our focus. Removing the initial national/country terms identified 120 studies, with no country level estimates. Several single hospital setting estimates exist for England and other countries, but the percentage of hospital-associated infections reported relies on identified cases in the absence of universal testing. Added value of this study
This study provides the first national-level estimates of all symptomatic hospital-acquired infections with SARS-CoV-2 in England up to the 31st July 2020. Using comprehensive data, we calculate how many infections would be unidentified and hence can generate a total burden, impossible from just notification data. Moreover, our burden estimates for onward transmission suggest the contribution of hospitals to the overall infection burden. Implications of all the available evidence
Large numbers of patients may become infected with SARS-CoV-2 in hospitals though only a small proportion of such infections are identified. Further work is needed to better understand how interventions can reduce such transmission and to better understand the contributions of hospital transmission to mortality.

## INTRODUCTION

The SARS-CoV-2 pandemic is a global public health priority.^1^ Based on experience with other highly pathogenic coronaviruses within-hospital transmission may readily occur without sufficient infection control and hospitals may play an important role in amplifying transmission.^2^ Moreover, many patients acquiring SARS-CoV-2 in hospitals are at high risk for severe outcomes and subsequent mortality.^3^ Quantifying hospital-acquired transmission of SARS-CoV-2 is thus important both for prioritising control efforts and for understanding the contribution of hospitals to sustaining the community epidemic.

SARS-CoV-2 transmission in healthcare settings has been reported in many countries.^3–6^ As the precise time of infection is rarely known, establishing whether an infection is hospital-acquired remains a challenge. For SARS-CoV-2, hospital-acquired infections are usually defined by comparing the time of admission and subsequent symptom onset^7^ or first positive test.^8^ If the delay is much longer than the incubation time, then it is likely that an infection is hospital-acquired. Thus, the proportion of patients with a hospital-acquired SARS-CoV-2 infection will depend on the definition used, with uncertainty driven by the unobservable nature of infection and the incubation period distribution. Records for all hospitals in England, using standard definitions and testing data, indicate that 15% of detected SARS-CoV-2 infections in hospitalised patients could be attributed to hospital-acquired transmission ^8^ with analysis of data from single NHS Trusts suggesting a similar level.^3,9^

In the absence of frequent universal testing of all inpatients, many hospital-acquired SARS-CoV-2 infections will not be identified by hospitals prior to discharge. Even with regular PCR testing of all inpatients regardless of symptoms we would expect to miss many infections because of short patient stays and potentially low PCR sensitivity 1-2 days after infection.^10^

In the spring of 2020 in England, the majority of inpatient testing only occurred in those with symptoms, either on admission or during hospital stay.^11^ Many patients who develop a symptomatic infection will do so after discharge (Figure 1) as hospital stays are typically shorter than the interval from infection to symptom onset (median length of stay = 2.4 days, standard deviation = 0.4 days, for non-COVID patients in England vs. incubation period average of 5·1 days ^12^). Thus, there may be a substantial proportion of infection and subsequent onward community transmission seeded by hospital-acquired infections which has been difficult to quantify. Additionally, a substantial proportion of infected individuals never progress to be symptomatic.^13^

**FIGURE 1:**
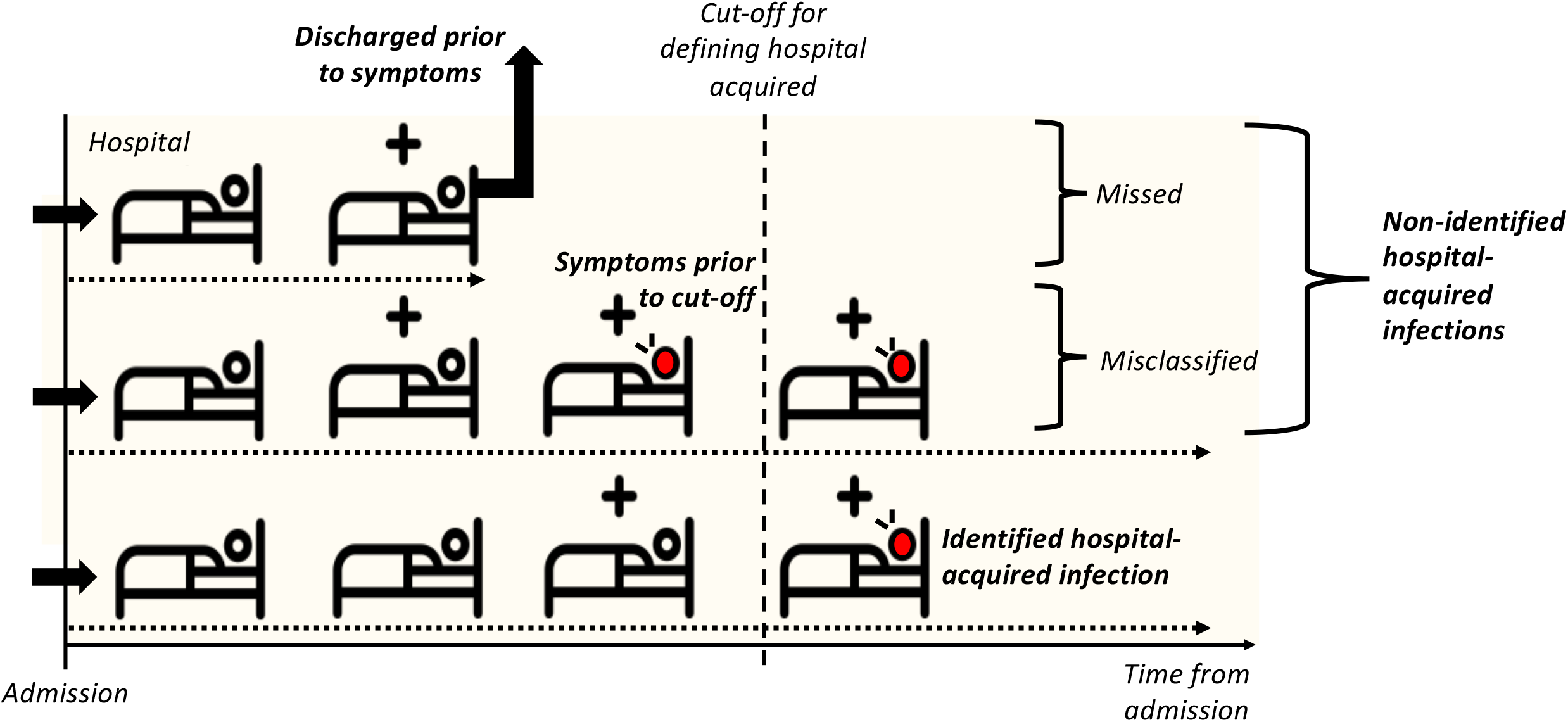
How might we underestimate hospital-acquired (HA) infections? With no asymptomatic screening in hospitals, detection of a hospital-acquired case relies on symptom onset prior to patient discharge. In the schematic a “+” above the bed denotes a hospital-acquired infection, and a red patient denotes one with symptoms. A patient with COVID-19 identified as being due to a hospital-acquired infection is one with symptom onset after a defined cut-off (e.g. >7 days from admission to symptom onset but prior to discharge, bottom row patient). Patients with unidentified hospital-acquired infections are those with a symptom onset after discharge (top row patient, “missed”) or those with symptom onset prior to the defined cut-off (middle row patient, “misclassified”). We focus on symptomatic infection: there will also be unidentified asymptomatic hospital-acquired infection which we do not include. We estimate that fewer than 1% of individuals with symptom onset >7 days from admission will have been infected in the community.

In this analysis, we used national, patient-level datasets of patients hospitalised with COVID-19 to estimate the contribution of hospital settings to the first wave of COVID-19 in acute Trusts in England. We estimated the proportion of symptomatic hospital-acquired infections that have not been identified as hospital-acquired and modelled onward transmission from these unidentified infections in the community. We hence quantified the likely contribution of hospital-acquired infections to the first wave of SARS-CoV-2 infections in England.

## METHODS

Our primary aim was to estimate the total number of symptomatic hospital-acquired SARS-CoV-2 infections in England from 1st January to 31st July 2020. For each identified hospital-acquired infection, we estimated how many were unidentified. Our secondary aim was to estimate the contribution of these unidentified hospital-acquired infections to the community epidemic.

All analyses were conducted in *R* version 4.0.3^14^ with code available on Github.^15^ The steps in the analysis are outlined in Figure 2.

**FIGURE 2:**
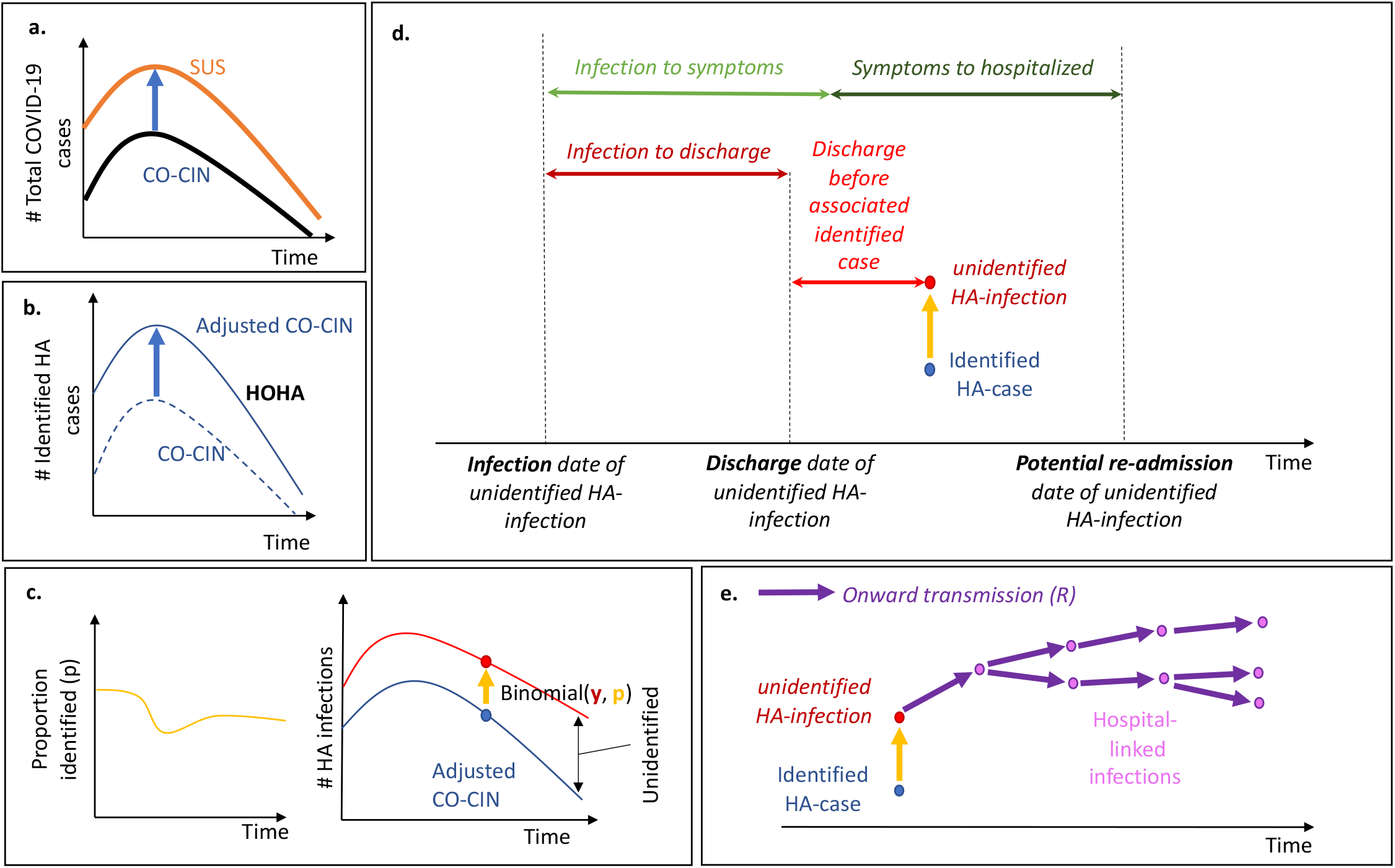
The analysis steps: (a) CO-CIN is inflated to match total COVID-19 hospitalised cases in SUS. (b) The same weekly adjustment is used to estimate the number of identified hospital-onset, hospital-acquired (HOHA) cases. (c) The length of stay for non-COVID-19 hospital patients and incubation period distribution is used to generate estimates of the proportion of hospital-acquired infections that would be identified (Figure 1). This proportion (*p*) is used to estimate how many unidentified hospital-acquired infections there would be for each identified hospital-onset hospital-acquired infection by assuming a Binomial distribution and calculating the number of “trials” or “unidentified” hospital-acquired infections there were. (d) The unidentified hospital-acquired infections with symptom onset after discharge (“missed”) may return to hospital as a COVID-19 case: the trajectory of their disease is calculated to determine their contribution to hospitalised cases. (e) These “missed” unidentified hospital-acquired infections are assumed to contribute to onward transmission in the community: here we capture four generations of transmission to estimate the number of hospital-linked infections and subsequent hospitalised cases under different *R* estimates.

### Data sources

We used two COVID-19 patient data sources (Supplementary 2). The first is the ISARIC4C UK COVID-19 Clinical Information Network (CO-CIN) study,^16^ using data collected before 3rd December 2020 but filtered for symptom onset before 1st August 2020. This is because CO-CIN is an enrollment-based study and individuals can be added retrospectively, up to several months after admission. CO-CIN is a national cohort of COVID-19 patients, representing approximately two thirds of COVID-19 UK admissions during the first wave of SARS-CoV-2 infection. Not all National Health Service (NHS) Trusts or sites are represented in the data as some have specialist roles that do not involve inpatient acute medical care: NHS Trusts are groupings of hospitals and other healthcare settings of which there are 223,^17,18^: our CO-CIN extract had data from 208 acute medical care Trusts. The study recorded admission date, discharge date, and date of symptom onset for patients. We excluded CO-CIN participants without a recorded admission and symptom onset date (Supplementary 2).

The second is the SUS dataset^19^ which contains data on all patient admissions and discharges for all Trusts in England. The SUS data were linked with testing data (Second Generation Surveillance System (SGSS))^19^ to derive length of stay distributions for non-COVID-19 patients and total COVID-19 hospital admissions by week and NHS Trust.

These two data sources have their respective strengths and limitations. The CO-CIN data include information on the date of symptom onset^20^ but is only a subset, albeit the majority, of all hospitalised COVID-19 patients, while the linked SUS/SGSS data include all known hospitalised COVID-19 patients but lack information on symptom onset date. Symptom onset dates do not rely on knowledge of testing regimens which vary over time and between Trusts. To address these different issues, we decided to use SUS data to adjust CO-CIN information to account for enrollment variation between settings, giving a database combining the best features of both.

### Setting

Our baseline population is all acute English Trusts in CO-CIN. Acute Trusts are defined as an NHS Trust with only acute hospital sites enrolled in CO-CIN (as opposed to Community or Mental Health facilities). These are aggregated as a single “England” population for our main analysis. A sensitivity analysis modelled the individual acute Trust level prior to aggregation (Supplementary 12).

### Length of stay distribution

We used empirical length of stay (LoS) estimates for non-COVID-19 patient stays from SUS for each English acute Trust in CO-CIN for patients admitted each week (Supplementary 2). To get a LoS distribution for England, LoS estimates across all including Trusts were pooled by week.

#### a. Identifying COVID-19 cases as infected in hospital

We used information on symptom onset and hospital admission from CO-CIN to estimate the number of hospital-acquired COVID-19 cases per day in each Trust. In our baseline analysis we defined an identified healthcare-acquired infection as an inpatient with symptoms onset more than 7 days after admission (Table 1) aligned with English definitions and the ECDC definition for a Probable (8-14) and Definite (>14days) healthcare-associated COVID-19 case^7,21^. In sensitivity analyses we explored cutoffs of 4 and 14 days.

**Table 1:**
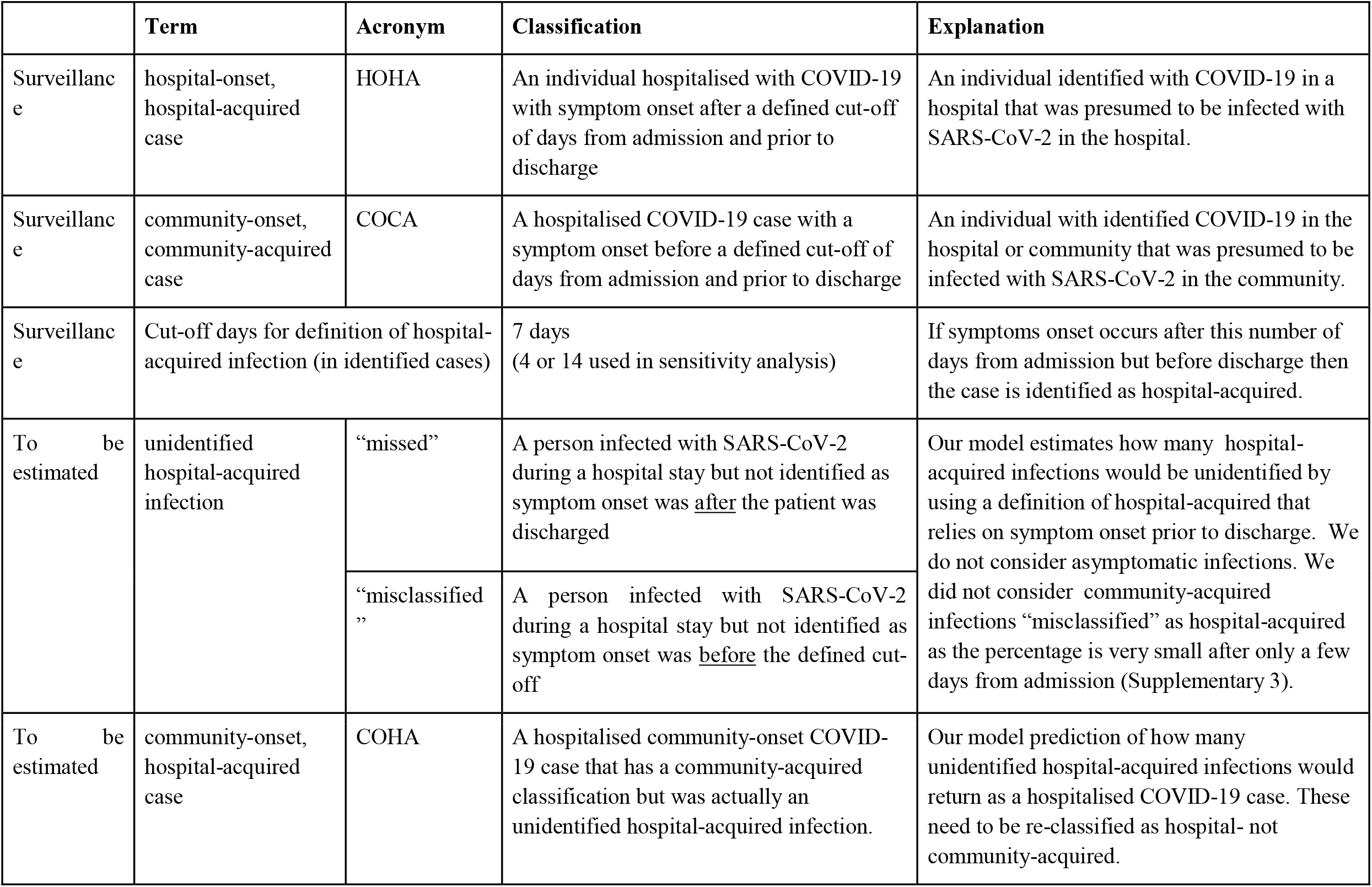
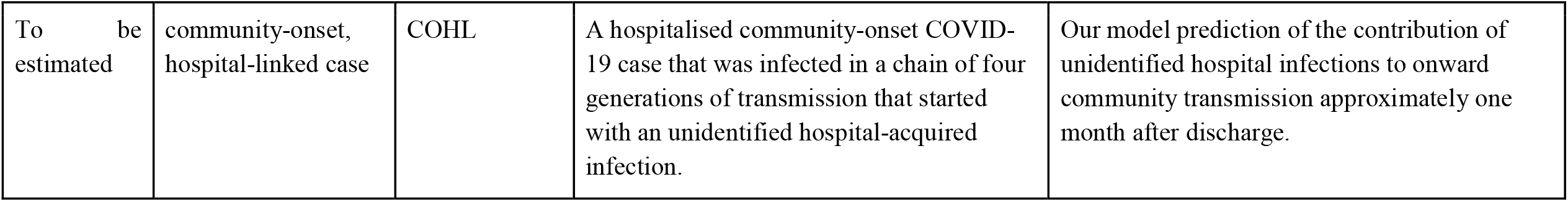
Case definitions. Terms are distinguished between surveillance definitions and quantities estimated in the analysis. Additional definitions are given in Supplementary 1.

#### b. Accounting for enrolment into CO-CIN

We calculated the proportion of COVID-19 patients recorded in SUS in a given week that were included in the corresponding CO-CIN data. We then weighted the weekly estimates of the number of hospital-acquired infections from the CO-CIN data using the inverse of these weekly proportions to obtain estimates of identified hospital-acquired COVID-19 cases corrected for under-reporting in CO-CIN (Supplementary 4) (assuming no bias in enrolment of hospital versus community-onset cases).

#### c. Proportion of hospital-acquired infections that are identified

To calculate the proportion of symptomatic hospital-acquired infections that were identified as such we considered the probability that a patient with a hospital-acquired infection has a symptom onset that falls in the definition period, i.e. before discharge and after the cut-off threshold (Figure 1). The calculations were based on the incubation period and length of stay distribution of non-COVID-19 patients and assumed that all infections led to a symptom onset: hence it is the proportion of hospital-acquired infected individuals that will ever have symptoms and are identified (Supplementary 5). Uncertainty was included by sampling from parameter distributions (Table 2, Supplementary 10).

**Table 2:**
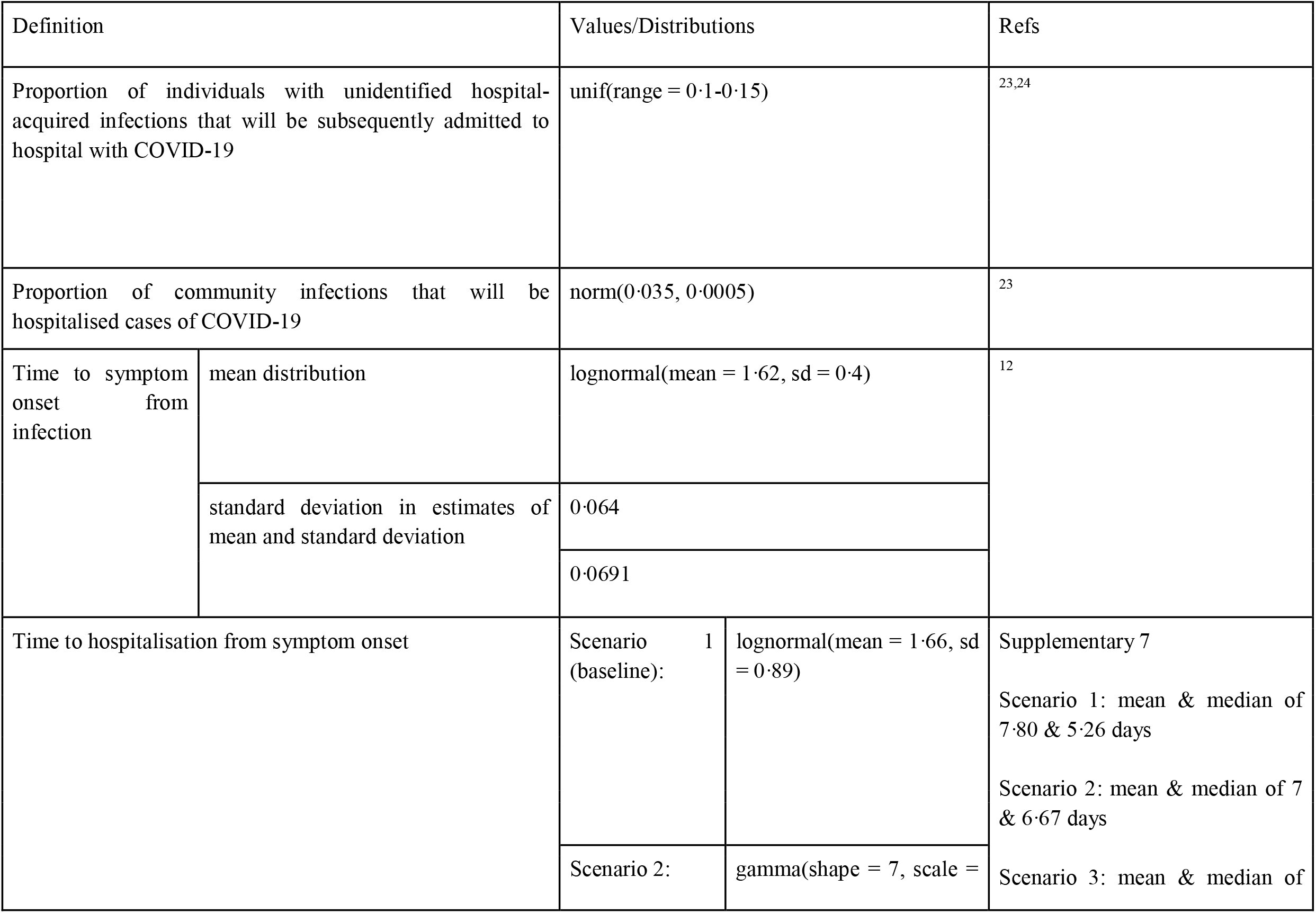
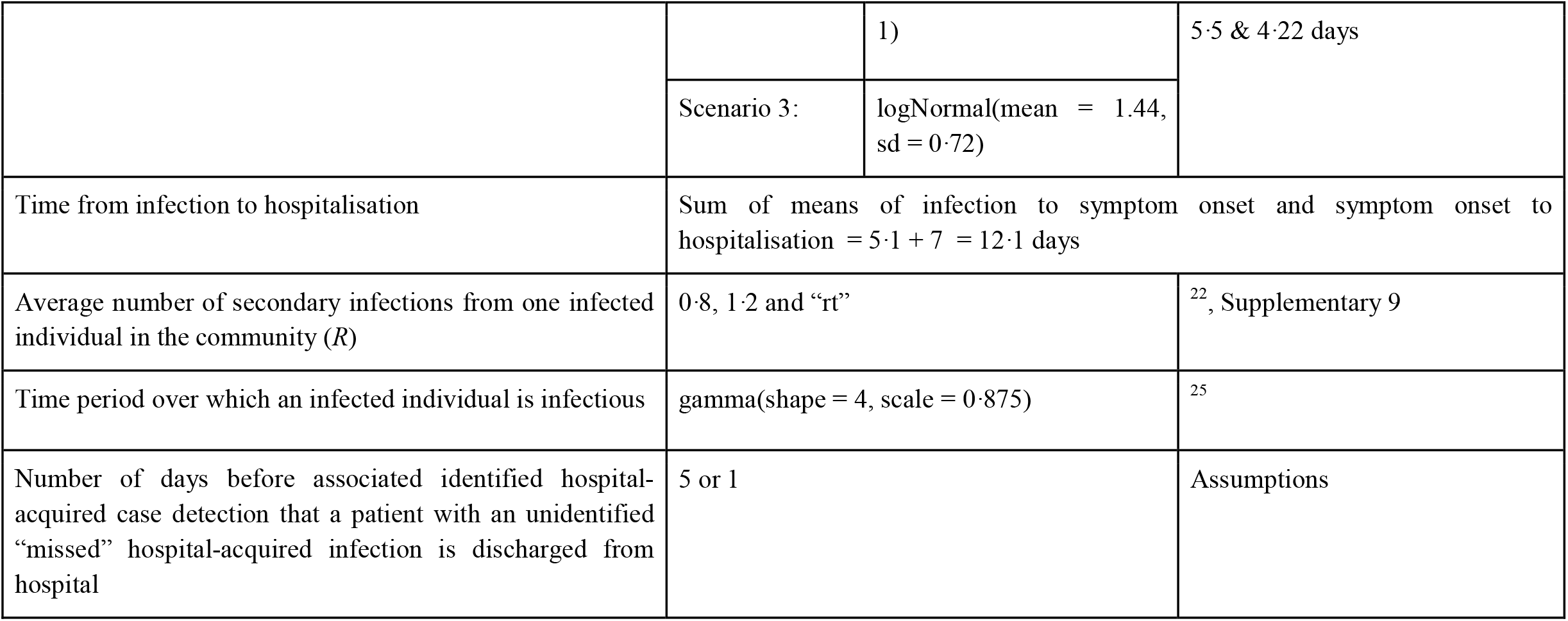
Parameters values used in the model. See Supplementary 6 for more details.

Patients with these unidentified hospital-acquired infections consist of patients with “missed” hospital-acquired infections where symptom onset occurs after discharge and patients with “misclassified” hospital-acquired infections where symptom onset occurs before discharge and before the definition cutoff days from admission (Figure 1, Table 1). We did not consider those who acquire infection but remain asymptomatic.

We estimated that fewer than 1% of inpatients with symptom onset 5 or more days after admission were latently infected when admitted (Supplementary 3). Hence, our definition of “misclassified” only considers those “hospital-acquired” infections misclassified as “community-acquired”.

#### d. Reclassifying community-acquired as hospital-acquired

To determine the contribution of unidentified hospital-acquired infections to the hospital burden, we estimated when an unidentified “missed’’ hospital-acquired infection would return as a hospital admission by generating the entire disease progression trajectory for each unidentified “missed” hospital-acquired infection (Figure 2).

We calculated the time from infection to discharge using the length of stay distribution of non-COVID patients (Supplementary 8). For the model generated patients with an unidentified infection, we assumed a date of discharge of 5 days before the detection date of the associated identified COVID-19 case (Figure 2c). This corresponds to the difference in the average length of stay of identified SARS-CoV-2 positive cases (∼7 days) and those thought to be SARS-CoV-2 negative (∼2 days) in SUS. We explore setting this to 1 day in the sensitivity analysis.

From this date of discharge, we calculated what proportion of these unidentified “missed” infections would have been expected to return as a hospitalised COVID-19 case and when, which varied for each simulation (Figure 2, Supplementary 6). Symptom onset is hard to define, hence we used a scenario analysis to explore three different distributions for the symptom onset to hospitalisation parameter (Table 2, Supplementary 7).

#### e. Hospital-linked cases

To account for onward transmission in the community from patients with unidentified “missed” hospital-acquired infections we estimated “hospital-linked infections’’: this time series was calculated by estimating four generations of onwards infection under varying assumptions about the reproduction number (Supplementary 6). This is approximately the number of infections caused within one month after discharge (∼6·7 day serial interval, Supplementary 6).

We explored three reproduction number values: a constant value of 0·8 or 1·2 with a range generated as +/- 5% of the constant value. For a time-varying estimate “*Rt”* we used upper/lower bounds for the 50% credible interval from a publicly available repository ^22^ (Supplementary 9).

## RESULTS

### Classified hospital-acquired cases

In CO-CIN, using a symptom onset-based definition, we found 7% (n = 65,028) of COVID-19 cases in acute English Trusts were classified as hospital-acquired (having a symptom onset 8 or more days after admission and before discharge) before 31st July 2020. By adjusting for enrolment in CO-CIN (Figure 2b), we estimated that with this same cut-off there were 6,640 “hospital-onset, hospital-acquired” identified cases across acute English Trusts up to the 31st July 2020.

### Proportion of infections identified

We estimated 30% (20-41%, range across weeks and sampling, Supplementary 10) of symptomatic hospital-acquired infections (using a 7 day cut-off) were identified using a symptom onset based definition for England. Across all acute English Trusts the range was 0-82% (Figure 3). The proportion identified decreased with increasing cut-off day from admission (Figure 3c). The estimates are highly sensitive to LoS distributions (Supplementary 2). These results imply that for every single identified hospital-acquired COVID-19 case (using a 7 day cut-off) there were, on average, two unidentified symptomatic hospital-acquired infections (Figures 1&2).

**FIGURE 3:**
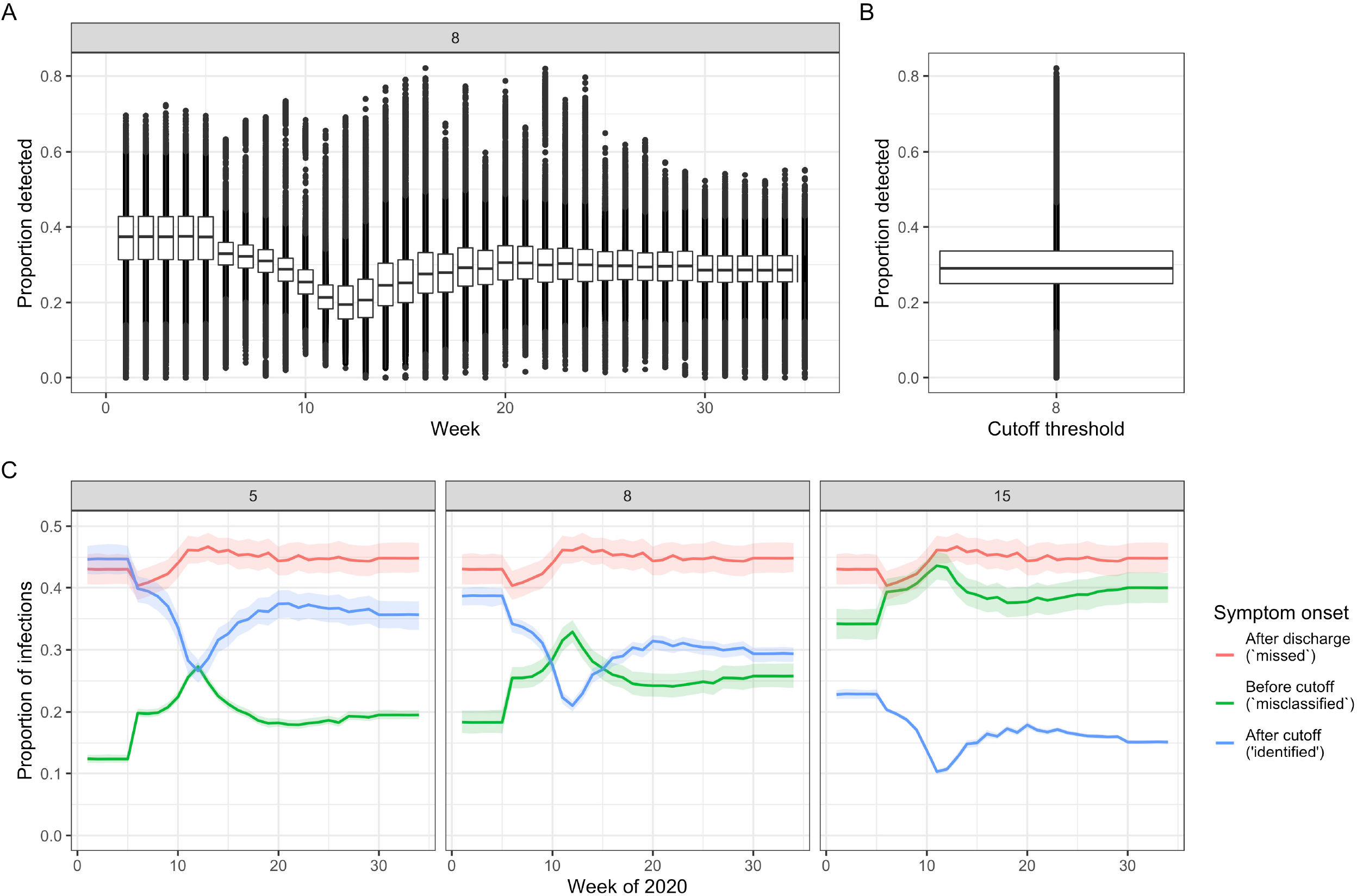
Proportion of symptomatic hospital-acquired infections identified, given by week (A) and over all weeks (B) at a 7 day cut-off, for all acute English Trusts. Each datapoint is the value from a single Trust for each of 200 samples. The boxplot highlights the median and 25th-75th quantile. (C) For England (the aggregate setting) the proportion of patients with hospital acquired infections split by those that are identified (blue) due to a symptom onset starting at a set number of days from admission (grey box) and before discharge, and those unidentified with symptom onset after discharge (“missed”, red) or before the cut-off (“misclassified”, green). The coloured lines represent the mean, and the shaded areas the 95% percentiles over the 200 samples.

### Contribution of missed infections

We estimated that across England, 20,000 (mean; 95% range over 200 simulations: 19,200, 21,100) hospital-acquired infections were unidentified from acute Trusts if a 7 day symptom-based cut-off was used to identify hospital-acquired cases. The majority of patients with unidentified hospital-acquired infections were not identified due to the discharge of the infected patient prior to symptom onset (“missed”) (Figure 1 and 3c): 12,300 (11,400, 13,400) in total.

A proportion of the patients with unidentified hospital-acquired infections that have symptom onset after discharge will return as hospitalised cases: we found 1,500 (1,200, 1,900) or 2·1% (1·7%, 2·6%) of cases originally classified as “community-onset, community-acquired” should be classified as “community-onset, hospital-acquired” for a 7 day cut-off.

We found that there could have been 47,400 (mean; 95% range over 600 simulations: 45,000, 50,000 for the time varying *R* value) infections of individuals in the community secondary to patients with unidentified infections acquired in the hospital which had symptom onset after discharge (“missed”) over the first wave. We estimated that these would result in 1,600 (1,600, 1,700) “community-onset, hospital-linked” cases with a 7 day cut-off. The values are reduced by one-third with an *R* constant at 0.8 (Supplementary 11). These contribute 2·3% (2·1%, 2·4%) of “community-onset, community-acquired” cases over the first wave with a 7 day cutoff and under both scenario 1 or 2 (Supplementary 11).

This contribution of community-linked cases to hospital admissions with COVID-19 varied depending on the timing of hospital admission post symptom onset (captured here by Scenarios 1-3, Table 2, Figure 4). The proportion of COVID-19 hospital admissions due to hospital-transmission was greatest when total case numbers first declined (peak in COHL in Figure 4D at ∼4% in late April).

**FIGURE 4:**
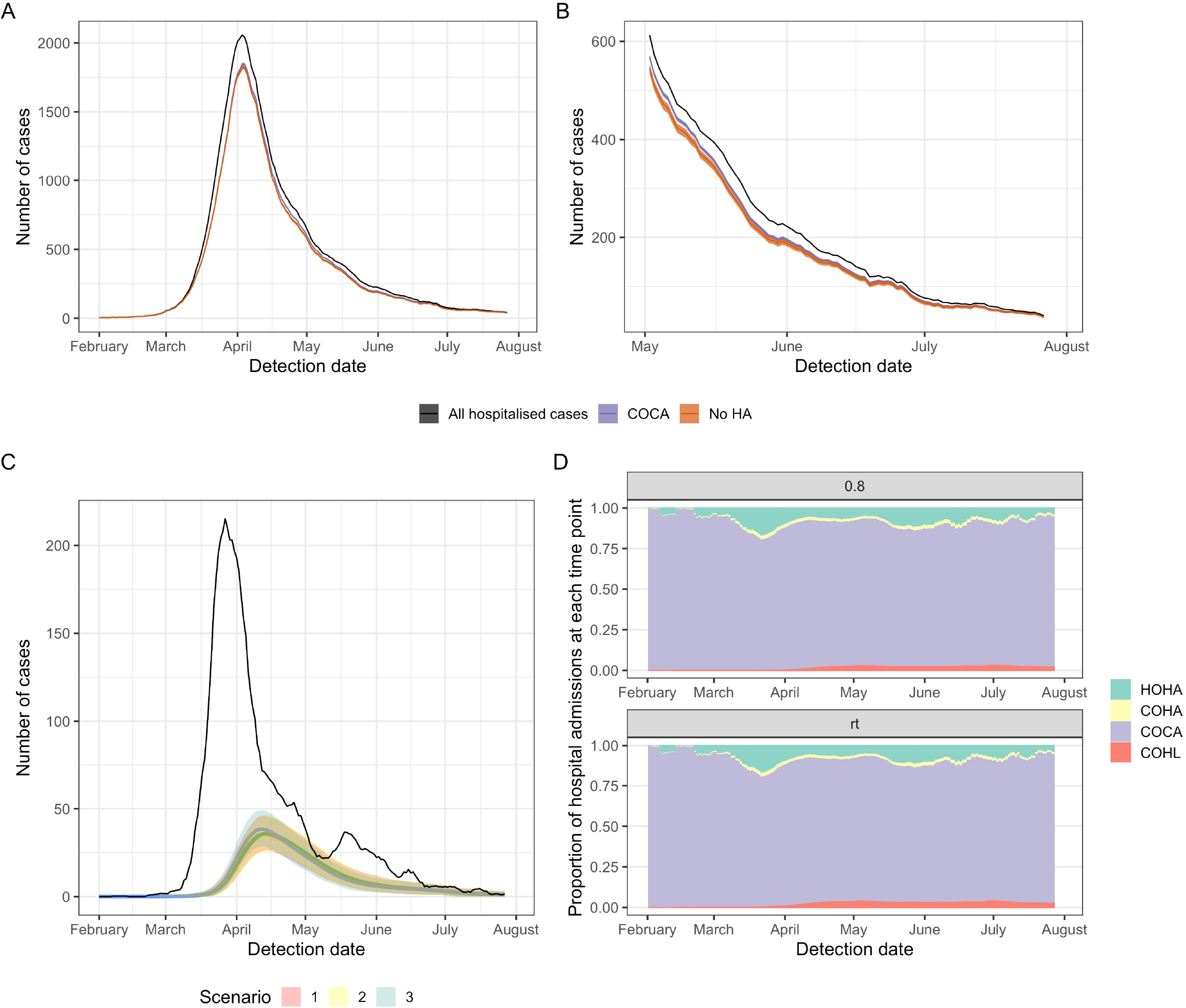
(A) Total COVID-19 admissions with model adjusted definitions from “community-onset, community-acquired” (COCA) for Scenario 1 for the whole study period (January - 31st July 2020) and (B) for the end of the study period (May - 31st July 2020). The counterfactual of no hospital transmission (“No HA”, orange) is compared to the adjusted model estimate of COCA (purple) and total admissions (black) for a time-varying R estimate. (C) The number of hospital-onset, hospital-acquired (HOHA) cases (black) is similar in magnitude to the number of community-onset hospital-linked (coloured lines, COHL) under the three scenarios for hospital admission after symptom onset. (D) The proportion of all hospital admissions in England that were estimated to be HOHA (green), community-onset, hospital-acquired (COHA, yellow), COCA (purple) and COHL (red) under two example R values (constant: 0·8 and time-varying “rt”) and Scenario 1. All outputs take a threshold cut-off value for defining hospital-acquired as a symptom onset more than 7 days from admission. All outputs are the rolling 7-day mean for the mean over 200 simulations with 5-95% ranges in shaded areas in (C).

The number of unidentified hospital-acquired infections and hence reclassification levels increased or decreased under a 14 or 4 day cutoff respectively (Supplementary 11).

### Contribution of hospital settings to cases, infections and onward transmission

To summarise, we estimated that there have been 26,600 (mean, 95% range over 200 simulations: 25,900, 27,700) hospital-acquired SARS-CoV-2 infections in acute English Trusts (E, Figure 5) with a 7 day cutoff. Of these, a total of 15,900 (15,200, 16,400) infections correspond to patients with COVID-19 that were identified as symptomatic cases in hospitals (B+C, Figure 5): as such 60% of hospital-acquired infections were identified. Over the whole first wave, 15% (14·1%, 15·8%) of cases classified as community-acquired were estimated to be hospital-acquired or hospital-linked ((C + F) / (A - B), Figure 5).

**FIGURE 5:**
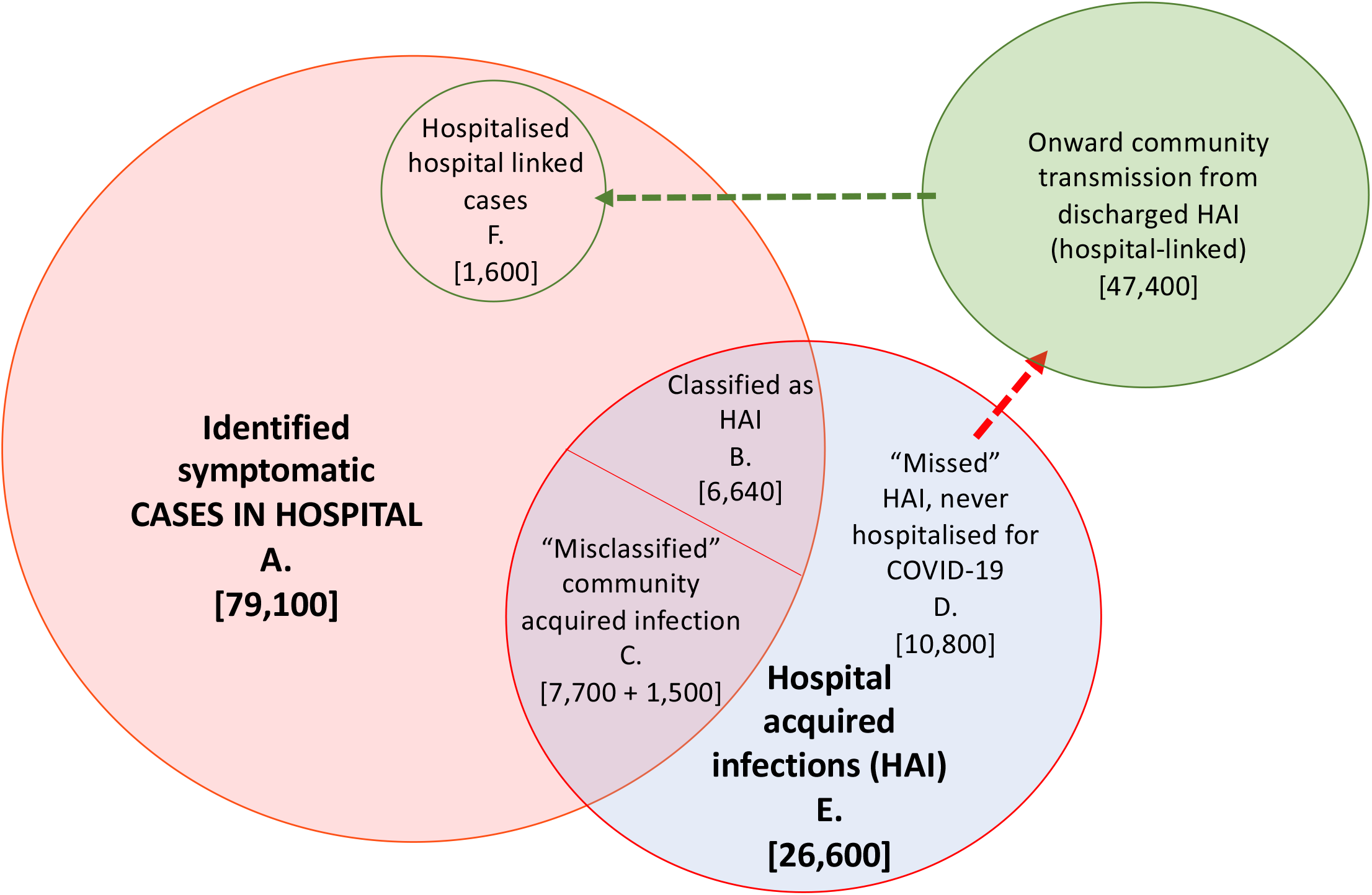
Summary figure of estimated values for patients with hospital-acquired symptomatic infections and onward community transmission with a 7 day cut-off for symptom onset after admission and prior to discharge for defining a patient with hospital-acquired infection. Note here that the “misclassified” (C) includes those “missed” unidentified infections that return to hospital later as a hospitalised COVID-19 case (1,500 “community-onset, hospital-acquired” cases).

The estimated percentage of identified COVID-19 cases in hospitals that were hospital-acquired is then 20·1% (19·2%, 20·7%) ((B + C)/ A, Fig. 5). Accounting for onward transmission from unidentified “missed” hospital-acquired infections, we estimated that 22·1% (21·2%, 22·9%) of hospitalised COVID-19 cases were hospital-acquired or hospital-linked ((B + C + F)/A, Figure 5) using the median time-varying *R* value.

If 20·1% of COVID-19 cases identified in hospitals were hospital-acquired then, assuming that 3% of symptomatic cases were hospitalised, we estimated that hospital-acquired infections likely contributed to fewer than 1% of infections of the overall English epidemic of COVID-19 in wave 1.

Assuming similar levels of hospital transmission in non-acute English trusts suggests approximately 31,100 (30,300, 32,400) symptomatic infections could have been caused in total by hospital-acquired transmission in England.

### Trust level and Sensitivity analysis

When aggregated, the results from the individual Trust level predicted a slightly higher proportion of cases to be hospital-acquired (25% vs 20%) (Supplementary 12). Varying the day of discharge of the unidentified “missed” infections had little impact on total case numbers, but did affect hospital-linked cases (Supplementary 11).

## DISCUSSION

We estimated that 20·1% (19·2%, 20·7%) of identified COVID-19 cases in hospitals were likely to have been hospital-acquired infections and that within-hospital transmission likely contributed directly to 26,600 (mean, 95% range over 200 simulations: 25,900, 27,700) symptomatic infections, and a further 47,400 (45,000, 50,000) hospital-linked infections. These results are based on a 7 day cutoff for symptom onset from admission and prior to discharge for defining an identified hospital-acquired case.

Despite these levels of infection, we estimated hospital transmission to patients caused fewer than 1% of all infections in England in the first wave. To some extent this reflects effective infection prevention within hospital settings with over 4 million non-COVID-19 patients being cared for in hospital settings during this period. However, the high proportion of hospital cases that were due to hospital-acquired infections is worrying as these are the most vulnerable members of our society and hence may have the most severe consequences. Our estimates are also an underestimate as they do not include the likely substantial proportion of infections that are asymptomatic.^13^

This is the first study to estimate the total number of hospital-acquired infections (not just the percentage of known cases that are hospital-acquired) and their wider contribution to community transmission. In particular, we found that the contribution of hospital-acquired infections to the epidemic likely varied over time, increasing in importance as community infections initially dropped, emphasising the need to determine where most infections are occurring at any one time during an epidemic.

Our results show that relying on symptom onset as a detection method for hospital-acquired SARS-CoV-2 may miss a substantial proportion (> 60%) of hospital-acquired infections. This is dependent on the length of stay for non-COVID admissions but suggests that in many settings estimates of the number of infections due to transmission in hospital settings will be substantial underestimates. This is particularly relevant for low-resource settings with short lengths of stay for non-COVID patients and which rely on symptom onset screening.

The alternative detection method is routine testing of patients, which will confirm symptomatic as well as detect pre-symptomatic and asymptomatic SARS-CoV-2 infections. However, even with screening on admission, symptomatic or not, and retesting 3 days after admission, a portion of infections will likely not be detected due to short lengths of stay. Our estimates of the proportion of hospital cases that are due to hospital-acquired infection are higher than those from an England wide study^8^ and those from single hospital settings in the UK.^3,9,26–28^, as we estimate all hospital-acquired infections whether identified or not. Our estimates of all infections are similar to previous modelling work using an SEIR model which estimates that nosocomial transmission was responsible for 20% (IQR 14·4, 27·1%) of infections in inpatients.^29^

Our work implies that it may be effective to screen patients upon hospital discharge to detect infection, or to quarantine hospital patients on discharge to prevent ongoing community transmission: we estimate this would detect approximately 40% of hospital-acquired infections that would become symptomatic (that would otherwise be “missed” in Figure 3c) and hence, dependent on test sensitivity by time from infection, up to 70% of hospital-acquired infections could be detected. The onward community transmission from these infections may be especially important as community prevalence of SARS-CoV-2 infection decreases.

Currently, much more routine screening and testing is implemented in English hospitals contributing to the detection of infections prior to symptom onset or discharge.^30^ However, screening will need to be conducted with high frequency to avoid missing those infected prior to discharge, or to screen on discharge.

Further work is needed to determine the precise risk of returning as a hospital case for those infected in hospitals. If our values (10-15%) are found to be conservative, then this percentage could increase substantially. If it were found to be higher, reflecting the poorer health of hospitalised patients and hence potentially increased susceptibility, then the proportion of hospital cases that are hospital-acquired could increase to 30-40%.

The interpretation of our results are limited by several simplifications. Firstly, we did not explicitly capture disease and hospital attendance variation by age. Future work could stratify our estimates to account for an older and more vulnerable hospital population. Secondly, we likely underestimated the total number of hospital-acquired infections as we modelled only those that progress to symptoms since these are the ones contributing directly to hospital burden. This decision was made as our definition of what was a hospital-acquired case was dependent on symptom onset and asymptomatic proportion estimates are highly variable ^13^. Thirdly, we assumed a fixed number of four generations for onward transmission in the community, and did not account for infections in healthcare workers, nor in the setting to which hospitalised patients were discharged to, such as long-term care facilities. The impact of onward transmission from hospital-acquired infections may be underestimated in this work since these settings may have high levels and large heterogeneity in onward transmission or overestimated if four generations is longer than the average chain from recently hospitalised individuals.

Finally, identification of hospital infection using CO-CIN relied on symptom onset date, which may be unreliably recorded potentially leading to bias in the patient population. While we cannot assess the biases, it is reasonable to expect that symptoms were recorded well in a clinical setting, and frequently (∼65,000 patients included). An alternative definition of hospital-acquired infection reliant on the date of first positive swab would have its own limitations: patients could enter with symptoms and not test positive until more than a week into their stay for example.^26^

Due to the delay from infection to symptom onset, hospital-acquired transmission of SARS-CoV-2 may be missed under common definitions of hospital-acquired. We estimated that nearly 20% of symptomatic COVID-19 patients in hospitals in England in the first wave acquired their infection in hospital settings. Whilst this is likely to have contributed little to the overall number of infections in England, the vulnerability of the hospital community means that this is an important area for further focus. Increased awareness and testing, especially of patients on discharge, as is now commonly in place, is needed to prevent hospitals becoming vehicles for SARS-CoV-2 transmission.

## Data Availability

References for all data and code used are available via a github repository (https://github.com/gwenknight/hai_first_wave.git). All data is available by application to the ISARIC4C UK COVID-19 Clinical Information Network (CO-CIN) study and to PHE for the SUS and Second Generation Surveillance System (SGSS) data.

