## Supplementary file for "The contribution of hospital-acquired infections to the COVID-19 epidemic in England in the first half of 2020"

**Table of Contents**


Supplementary 1: Definitions
Supplementary 2: Datasets
- Trust and case number differences
- LoS distributions
Supplementary 3: Admission with infection levels
Supplementary 4: Comparing COCIN and SUS by week
Supplementary 5: Calculations of proportion undetected hospital-acquired SARS-CoV-2 infection
Supplementary 6: Parameterisation and additional methods
Supplementary 7: Symptom onset to hospitalisation
Supplementary 8: Infection to discharge calculations
Supplementary 9: *Rt* estimates
Supplementary 10: Uncertainty inclusion
Supplementary 11: Additional results
Supplementary 12: Grouped Trust level analysis

#### Supplementary 1: Definitions

| Term | Definition | Specifics for this analysis |
|---|---|---|
| **Case** | An individual that has COVID-19 (the disease due to SARS-CoV-2 infection) | |
| **Patient with a hospital-acquired SARS-CoV-2 infection** | A patient with an infection acquired in the hospital setting, whether identified or not | |
| **Identified hospital acquired infection** | An individual with a SARS-CoV-2 infection that has been identified as hospital-acquired | In this work SARS-CoV-2 infection is detected by a case with symptom onset prior to 5, 8 or 15 days from admission in line with the ECDC definition (1) |
| **unidentified hospital acquired infection** | An individual with a SARS-CoV-2 infection that has not been identified as hospital-acquired | Some of these will be misclassified as community-acquired, some will be "missed" as the patient is discharged before symptom onset. |
| **Symptom onset** | The self-reported start date of COVID-19 symptoms | Here we mostly use the CO-CIN data which has a symptom onset defined by the ISARIC protocol. |
| **Community-acquired** | A patient with an infection with SARS-CoV-2 that is classified as being acquired outside of the hospital in the community setting | Individuals with a symptom onset before the cutoff date, including before admission, are classified as community-acquired in CO-CIN. |
| **Hospital-linked** | A patient with an infection that was acquired by transmission in the community from a four-generation chain of transmission originating with a unidentified "missed" hospital-acquired infection | We assume that every hospital-acquired infection that is "missed" is discharged into the community and can cause onward transmission. We calculated the number over approximately one month after discharge (4 x 6.7 days). |
| **Classified** | The assignation of "community-acquired" or "hospital-acquired" to the infection within a hospitalised patient with COVID-19 | We use this to specify the current classification of a symptomatic infection. Hence a case could be classified as "community-acquired" but actually be "hospital-acquired". We chose to use classified as well as "identified" as some hospital acquired infections would not have been classified whilst some would. |
| **Identified** | The detection of hospital-acquired infection | |
| **Detection date** | the most recent of (1) date of symptom onset or (2) date of admission if this occurred after symptom onset for a patient with COVID-19, censored at date of discharge | For any "community-onset" case this was their admission date. For "hospital-onset, hospital-acquired" cases this was their date of symptom onset (Table 1). |

**Table S1: Common definitions**

**Supplementary 2: Datasets**

**Trust and case number differences**

For COCIN, we included 123 Trusts and 3 super-Trusts in the final data analysis (see Supplementary 4 for definition of super-Trusts, basically pooled Trusts to account for frequent transfers).

SUS covers 589 Trusts in England. 319 of these reported a total of 91,319 COVID-19 cases up to 31st July 2020. 13,415 of these cases were not included in COCIN: suggesting that COCIN has a coverage of ~85% of the total.

**CO-CIN data inclusion**

Using the 3rd December CO-CIN data extraction, there were 104,672 unique subject IDs. Of these 78% had a symptom onset and admission date. 62%, or 65,028/104,672 unique subject IDs were included in the final dataset. The included cases were those with (i) a symptom onset date, (ii) an admission date, (iii) a symptom onset date after the 12st January 2020 and (iv) a symptom onset date before the 31st July 2020. Most patients had a symptom onset before admission (Figure S1).

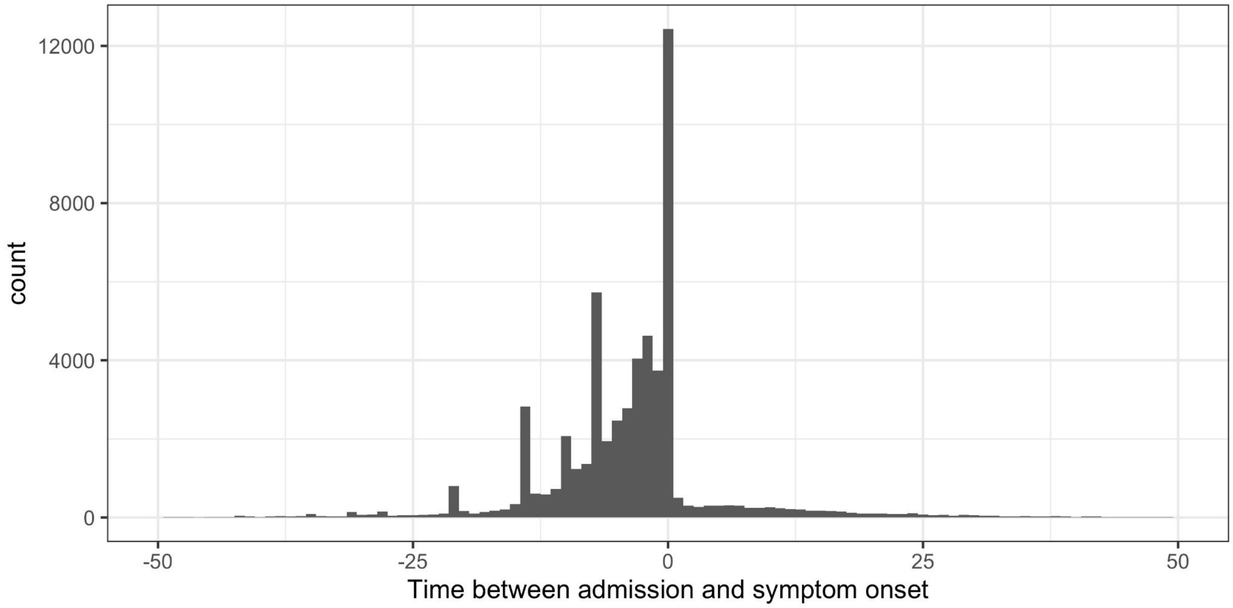


**Figure S1: Data from CO-CIN on time between admission to hospital and symptom onset.**

We defined a date of "detection" as the most recent of (1) date of symptom onset or (2) date of admission if this occurred after symptom onset for a patient with COVID-19, censored at date of discharge. For any "community-onset" case this was their admission date. For "hospital-onset, hospital-acquired" cases this was their date of symptom onset (Table 1).

**LoS distributions**

The length of stay (LoS) for non-COVID-19 positive patients is shown by week (in Figure S2) and over time (in Figure S3). Non-COVID-19 patients were defined as in-patients who never had a positive test, or who tested positive either after their hospital stay or more than 14 days before admission. Only one Trust (RX3) was removed as there were only 11 LoS data points (vs. a mean of 927 data points across other included acute Trusts) for non-COVID patients.

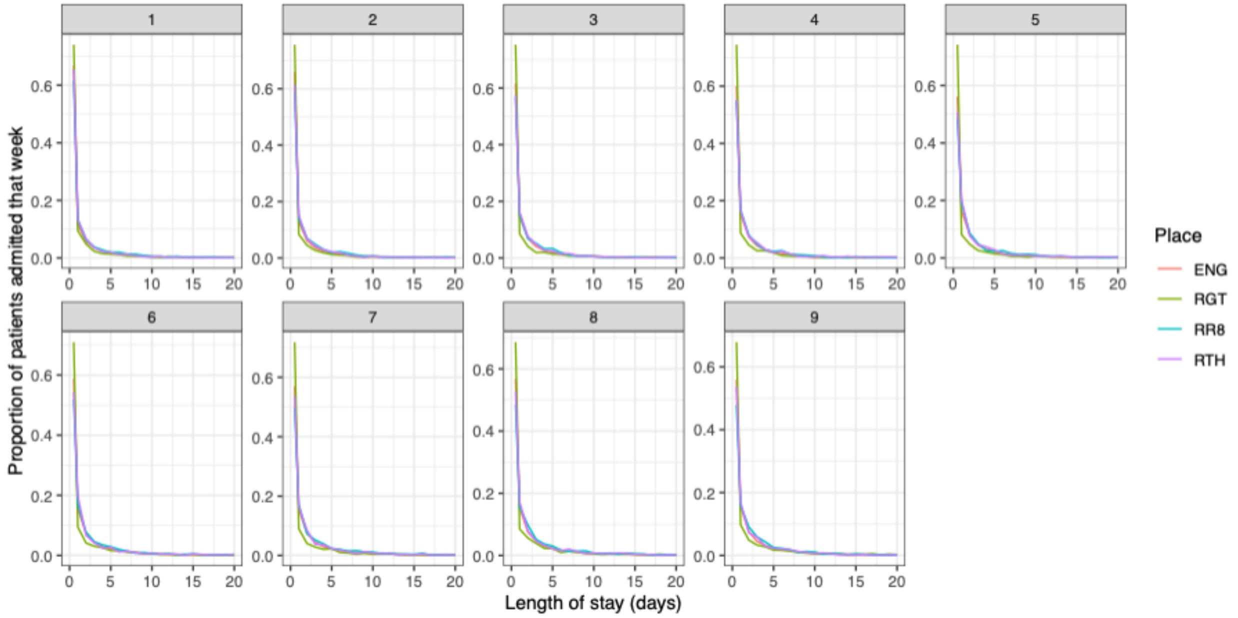


***Figure S2: Length of stay variation by week (facet) and three example Trusts (colour)***

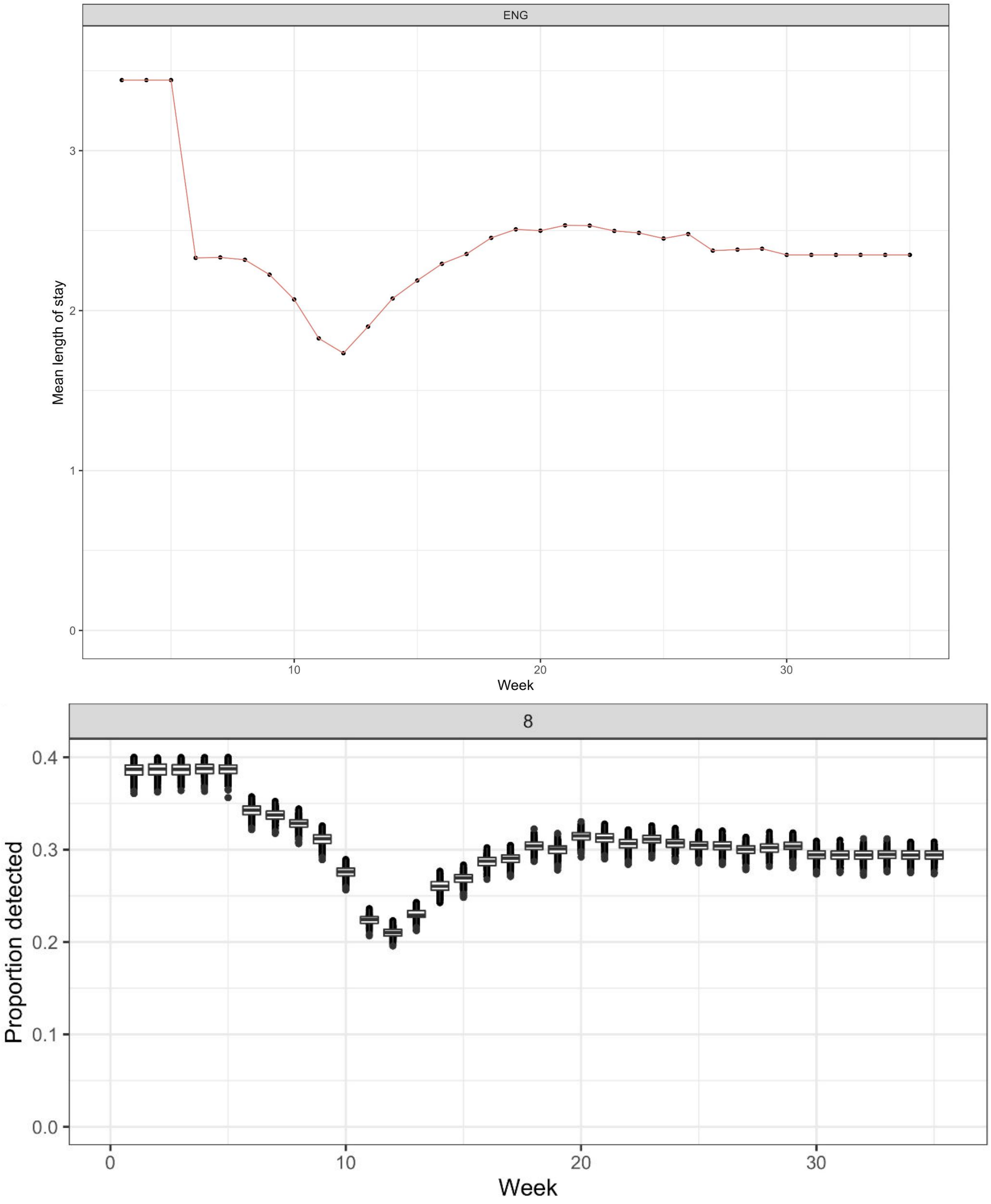


***Figure S3: Average length of stay over time for England in top panel to compare to proportion identified for England (equivalent to Figure 3A from main text, bottom panel)).***

**Supplementary 3: Admission with infection levels**

**What proportion of hospitalised patients with symptom onset after the cut-off day T had been infected in the community and admitted to hospital for a non-covid reason while latently infected?**

**Data**

The maximum prevalence of infection from seroprevalence surveys in the UK prior to September 2020 has been approximately:

0.5% from ONS (modelled, smoothed)(2)
0.3% from REACT1(3)

Between the 27$^{th}$ April & 10$^{th}$ May, ONS estimated prevalence of infection to be: 0.27 (0.17-0.41)%.

**Model**

The percentage of people at day T with COVID that acquired it in the community =

Prevalence of infection at entry x probability still in hospital at day T x probability symptoms developed after day T = (prev * (1-pexp(*T*,1/los)) * (1-plnorm(*T*,1.621, 0.418))*100.

Baseline measures

For example, using the ONS data for early May:

0.0027 * (1-pexp(*T*,1/los)) * (1-plnorm(*T*,1.621, 0.418))*100

For $T > 10$ this is zero due to very few patients remaining in hospital past this point (even assuming los for non-COVID of 7 days, which is an overestimate).

For $T = 5$, the value is 0.03 (0.02,0.04)%, 0.05 (0.03,0.08)% 0.07 (0.04,0.1)% for mean length of stays of 3, 5 or 7 days respectively. In conclusion < 0.1% of cases past day 5 are likely to be acquired in the community currently.

At the maximum prevalence:

At maximum prevalence 0.0054 * (1-pexp(*T*,1/los)) * (1-plnorm(*T*,1.621, 0.418))*100

For $T > 10$ this is zero due to very few patients remaining in hospital past this point (even assuming los for non-COVID of 7 days, which is an overestimate).

For $T = 5$, the value is 0.05 (0.04,0.07)%, 0.1 (0.08,0.13)% 0.14 (0.1,0.17)% for mean length of stays of 3, 5 or 7 days respectively. In conclusion < 0.2% of cases past day 5 are likely to be acquired in the community currently.

**Conclusion:** The prevalence was likely to be higher at the peak of the epidemic, but even at 10x higher this would be less than 1% of cases past day 5 being attributable to non-recent hospital transmission.

**Supplementary 4: Comparing COCIN and SUS by week**

There are several discrepancies between the Trusts enrolled in COCIN and SUS. The steps to calculate how to go from non-complete enrolment in CO-CIN to SUS (national COVID-19 case total data) are given below.

For each Trust in CO-CIN and each week (aggregated using lubridate::week (Grolemund and Wickham 2011)), the proportion of CO-CIN cases in SUS was calculated.

*When the proportion of SUS in CO-CIN was less than 1 (expected as CO-CIN enrolment based)*
The algorithm for a single Trust or England, for a set cutoff was

(1) Calculate the weekly proportion of CO-CIN cases in SUS
(2) Inverse this weekly proportion to give a multiplier
(3) In the cleaned (removed those with no subject onset or admission date), one row per subject CO-CIN, enter the multiplier for the week of the admission date for each subject
(4) Multiply each single hospital-acquired defined case by the multiplier for their week of admission to inflate the hospital-acquired case numbers. These were rounded to the nearest number.
(5) Aggregate over individual case data to get total number of
    (a) hospital-acquired cases (by summing over the inflated case numbers at the individual level)
    (b) Total cases (by summing over the multipliers: each single entry needs inflating)

Code in: trust_number_noso_all.R in https://github.com/gwenknight/hai_first_wave.git (4).

*When the proportion of SUS in CO-CIN was greater than 1 (unexpected as SUS should have all cases)*
If this proportion was greater than 1 (i.e. unexpected more cases in CO-CIN than SUS), then we explored the actual numerical difference in case numbers that was seen. If this difference in numbers was greater than 20% of the original total numbers in CO-CIN then we explored the difference further: 20 Trusts. The idea here is that especially in May / June there is a small number of cases admitted per week (< 5). It may be that a proportion >1 is then 2 in CO-CIN but only 1 in SUS. If their relative difference is not so big (< 20%) of the original CO-CIN data then we ignore this issue and set the proportion to 1.

For those to be explored further, we looked at the impact of capping the proportion at 1 and multiplying through the CO-CIN data to match the SUS data. If the total number of cases was greater than 150% of SUS then explored these further: this was the case for 5 Trusts.

In closer investigation we found that several of these Trusts had frequent transfers with other Trusts, for example three Trusts in one county, meaning that cases may be differently labelled as being in one Trust or the other in COCIN and SUS. This may be as SUS is based on test date and COCIN on symptom onset which may occur for a patient in different Trusts. To tackle this we aggregated Trusts with frequent transfers into super-Trusts. This results in three super-Trusts (R13, RR0, ESX) which included 2 (RT3, R1K), 2 (RRF, 02H), or 3 (RDD, RQ8, RAJ) Trusts and covered four of these problem Trusts. The fifth Trust (RBA) we removed from analysis as the discrepancy was substantial: more than 20 cases in COCIN than SUS at the peak and a secondary SUS peak that was not present in CO-CIN.

The resulting proportion of CO-CIN cases in SUS over time is shown in Figure S4.

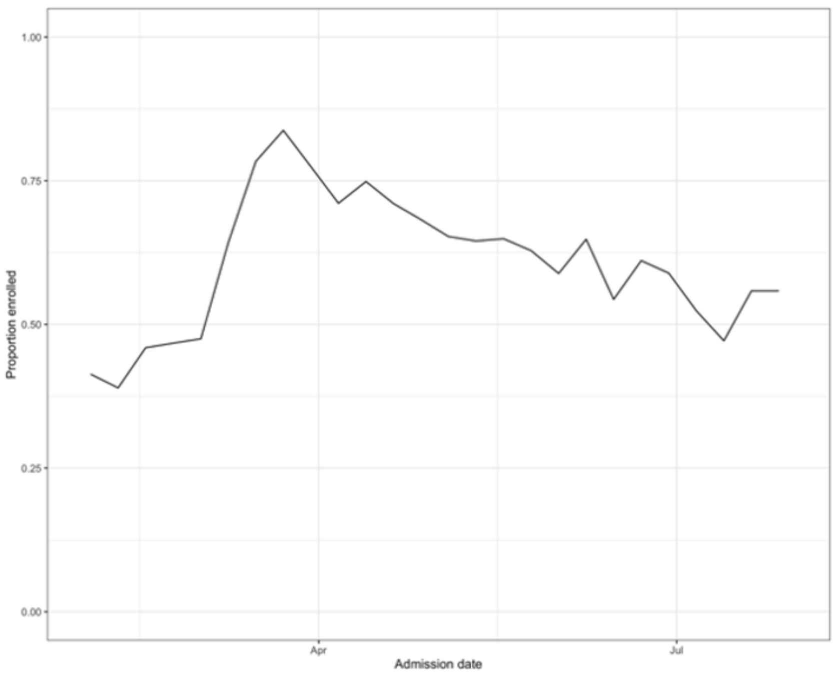


*Figure S4: Proportion of CO-CIN cases in SUS over time for acute English Trusts*

### Supplementary 5: Calculations for the proportion of undetected hospital-acquired SARS-CoV-2 infections

Hospital-acquired infections are here defined as patients who have symptom onset after a certain cut-off value $X$ after hospital admission. In particular, if $T_{\text{inf}}$ is the time of infection, $T_{\text{inc}}$ is the time from infection till symptom onset and $X$ is number of days after hospital admission of a hospitalised patient, then the patient is classified/detected as a nosocomial case if $T_{\text{inf}} + T_{\text{inc}} \geq X$. Only a subset of all hospital-acquired infections will be detected by this method. We estimated the proportion of hospital-acquired cases that get detected in the hospital based on information available from the CO-CIN and SUS data set. From that we could deduce the proportion of hospital-acquired infections that would be *missed* by this method. We assumed that the cut-off value $X$ is chosen large enough such that community-acquired cases can be excluded.

We implemented R functions for the calculations of the proportions of missed hospital-acquired infections based on the theoretical calculations below. The full code is available from: https://github.com/tm-pham/covid-19_nosocomialdetection.

#### CO-CIN Analysis

CO-CIN includes information on date of symptom onset of hospitalised patients. Let $LoS$ be the random variable representing the length of stay of hospitalised (non-COVID-19) patients and estimated from empirical data from SUS. Three types of hospital-acquired cases can be distinguished:

1. Patients with symptom onset before the cut-off $X$ days after admission, i.e. $\{T_{\text{inf}} + T_{\text{inc}} < X\}$
2. Patients with a symptom onset after discharge, i.e. $\{T_{\text{inf}} + T_{\text{inc}} > \text{LoS}\}$
3. Patients with a symptom onset after $X$ days after admission but before discharge, and with a length of stay of at least $X$ days, i.e. $\{T_{\text{inf}} + T_{\text{inc}} \geq X\} \cap \{\text{LoS} \geq T_{\text{inf}} + T_{\text{inc}}\}$

Only the last category of hospitsal-acquired cases will be detected by the method described above. On a given day, the probability that a hospital-acquired case is detected (using a cut-off of $X$ days) is given by

$$
\begin{aligned}
&P(\text{randomly selected patient is detected on a given day} \,|\, \text{patient is a nosocomial case}) && (1)\\
&= P(\text{randomly selected patient fulfills 3.}) && (2)\\
&= P(X \leq T_{\text{inf}} + T_{\text{inc}} \leq LoS) && (3)\\
&= \sum_{l=X}^{\infty} p_l P(X \leq T_{\text{inf}} + T_{\text{inc}} \leq l) && (4)\\
&= \sum_{l=X}^{\infty} \sum_{t=1}^{l-1} p_l P(X \leq t + T_{\text{inc}} \leq l) \cdot P(T_{\text{inf}} = t) && (5)\\
&= \sum_{l=X}^{\infty} \sum_{t=1}^{l-1} p_l P(X \leq t + T_{\text{inc}} \leq l) \cdot \frac{1}{l} && (6)\\
&= \sum_{l=X}^{\infty} \sum_{t=1}^{l-1} \frac{p_l}{l} P(X - t \leq T_{\text{inc}} \leq l - t) && (7)
\end{aligned}
$$

We adjusted for the fact that over a given period of time, patients with longer length of stays are more likely to be encountered and to be infected in the hospital than patients with short length of stays. Hence, the probability that on a given day, a randomly selected hospital-acquired case has LoS $= l$ is given by

$$p_l = \frac{P(\text{LoS} = l) \cdot l}{\sum_{l=1}^{\infty} l \cdot P(\text{LoS} = l)}$$

We, further, assumed a constant force of infection on each day, i.e., a non-COVID-19 patient is equally likely to get infected on each day and therefore $P(T_{\text{inf}} = t) = \frac{1}{l}$. This assumption was verified using data from Oxford (see below).

#### Probability of infection per day

In the above calculation, we assumed that a non-COVID-19 patient is equally likely to get infected on each day and therefore $P(T_{\text{inf}} = t) = \frac{1}{l}$. This assumption was based on hospital data from Oxfordshire. We fitted a generalised additive model with the probability of being tested positive for SARS-CoV-2 dependent on the day of hospitalisation accounting for age, gender, ward type, and ethnicity, using a logit link.

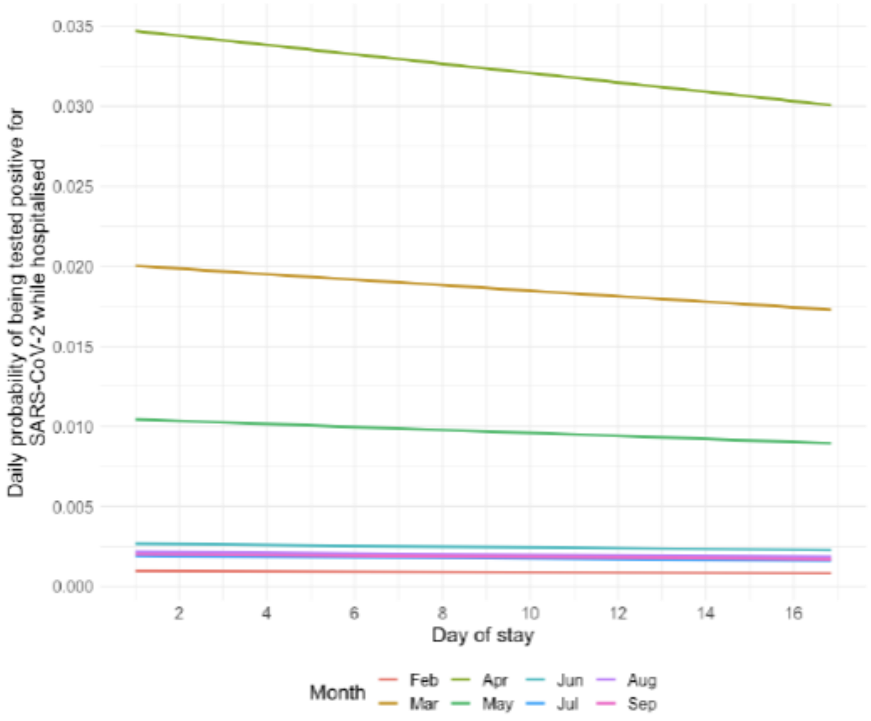


**Figure 1.** Daily probability of being tested positive for SARS-CoV-2 while hospitalised

**Supplementary 6: Parameterisation and additional methods**

| Parameter in R code | Definition | Literature | Notes | Base case |
|---|---|---|---|---|
| *prop_miss_hosp* | Proportion of recently hospitalised patients with missed hospital-acquired infections that will be subsequently admitted to hospital with COVID-19 | Infection hospitalisation ratio that ranged from < 5% in those aged 40 to > 40% in those aged 80+ (5)<br>3,4,8,12,17,18% infections are hospitalised for 10yr age groups from 30 to 80+ respectively (Table 3, (6))<br>Non-COVID hospital population composed of 33% older than 70, 60% older than 50 (5 yr age group data used) (7) | Multiplying the proportion of the non-COVID hospital population in each age group by the risks in Knock et. al. leads to an upper estimate of 15%. These patients have previously been hospitalised so have a higher risk of re-infection than others in their same age group.<br>We assumed a uniform distribution between 10% and 15%. For each patient a Bernouilli trial then used this sample to assess whether the patient would return | unif(0.1,0.15) |
| *prop_comm_hosp* | Proportion of community infections that will be hosp. cases of COVID-19 | 3.5% (95% CrI 3.3%-3.7%) of people infected needed hospitalisation (5) | Assume normal distribution with mean from literature, and estimated standard deviation to match range | norm(0.035, 0.0005) |
| *time_inf_to_symp_mean* | Time to symptom onset from infection | Incubation period: mean of 5.1 days (8) | Use the Lauer distribution for analysis | 1.62 |
| *time_inf_to_symp_sd* | | Log normally distribution, with mean of 5.8 (95% CI 5.0 to 6.7) days (9) | | 0.4 |
| *time_inf_to_symp_mean_sd* | Standard deviation in estimates of mean and standard deviation time to symptom onset from infection | Incubation period: mean of 5.1 days (8) | Taken from range from Lauer et al | 0.064 |
| *time_inf_to_symp_sd_sd* | | | | 0.0691 |
| *time_symp_to_hosp_meanlog* | Time to hospitalisation from symptom onset | Gamma distribution, shape parameter equal to the mean of 7 days (standard deviation 2.65) " (10)<br>Analysis of "first" wave CO-CIN gives a mean of 7.7 days, median = 6 days and a range of 1-129.<br>Analysis of first few 100 cases in the UK (11) | Scenario (1) : Log normal distribution fitted to CO-CIN data (mean of 7 days, median 6 days)<br>Scenario (2): Gamma distribution as in Davies et al (gamma(7,1))<br>Scenario (3): Log-normal distribution from FF100 data (log-normal(1.44, 0.72))<br>(See below) | 1.66 |
| *time_symp_to_hosp_sdlog* | | | | 0.89 |

| | | | | |
|---|---|---|---|---|
| Time from symptom onset to hospitalisation | | Approximately 2 weeks | Sum of means = 7 + 5.1 = 12.1 days | |
| *R* | Average number of secondary infections from one infected individual in the community | Use the time varying estimates of *R* from *epiforecasts.io* as well as constants | Constant or time varying estimate | 0.8, 1.2 and “rt” |
| *infectious_shape* | Time period over which an infected individual is infectious (time from exposure to infection) | Duration of clinical infectiousness: gamma (shape=3.5, scale = 4) (10) | Taken to be as in Davies et al as a balance between underestimating due to lack of pre-clinical period vs. an overestimate if look at serial interval estimates (3 - 6 days (12)) | 4 |
| *infectious_scale* | | Generation time estimates: 3.95 to 5.20 days (12) | | 0.875 (3.5/4) |
| *cut-off_date* | Days from admission cut off for defining hospital-acquired case | Assumed 5, 8, 10, 14 | Will affect time series of hospital-acquired cases | 5 |

*Table S2:* Parameters values used in the model. Extended version of Table 2.

**Serial interval:**
Latency period mean of 5.1 days
Infectious period mean of 3.4 days
Subsequent infection mean of 5.1 + uniform(0,1) * 3.4 = mean of 6.8 days

For each infection, a latency period, infectious period and uniform random number were sampled. An "*R*" number of subsequent infections were then generated at a time latency period plus the uniform random number times the infectious period.

We chose to look at approximately the first month of transmission after discharge to limit the number of onward cases. It is likely that chains of transmission are short: 4 generations in China (13), and suggested to be short from genomic data in the UK and New Zealand (14,15).

**Additional methods**

Extending the methods given in the main paper we include further details for some of the stages in Figure 2 below.

**c. Proportion of hospital-acquired infections that are identified**

To calculate this we assumed that the daily risk of infection did not change with the day of hospital stay, supported by data analysis (Supplementary 5). The proportions of true hospital-acquired infections which are identified is dependent on (i) the assumed cut-off threshold and (ii) the length of stay (LoS) distribution for patients hospitalised for reasons other than COVID-19 and hence at risk of becoming infected, with the latter varying by week and setting.

**d. Reclassifying community-acquired as hospital-acquired**

To determine the contribution of unidentified hospital-acquired infections to hospitalised patient burden, we estimated when an unidentified "missed" hospital-acquired infection would return as a hospital admission by generating the entire disease progression trajectory for each unidentified "missed" hospital-acquired infection (Figure 2).

For the disease progression trajectory, the proportion returning to hospital was sampled using a Bernouilli trial and varied for each simulation (Table 2). For each individual that was expected to become a hospitalised case we sampled a time (i) from infection until discharge (ii) from infection to symptoms and (iii) from symptoms to potential hospitalisation (Figure 2, Table 2). The time since infection was subtracted from the time to hospitalisation (the sum of time to symptoms from infection and time from symptoms to hospitalisation) to calculate the time at which the unidentified "missed" hospital-acquired infected individuals would be identified but currently misclassified as a "community" case at hospital admission (new "community onset, hospital-acquired" cases, Figure 2, Table 1).

**e. Hospital-linked cases**

To account for onward transmission in the community from patients with unidentified "missed" hospital-acquired infections (due to symptom onset after discharge) we estimated "hospital-linked infections": calculated as first-, second-, third- and fourth-generation infections. This is approximately the number of infections caused within one month after discharge (~6.7 day serial interval, Supplementary 6) and assumes that most onward transmission chains are relatively short (13–15).

The time series for these was calculated by sampling a certain time to infection (a sum of a sample from the latency distribution and a sample from a uniform distribution on 0-1 multiplied by a sample from the distribution for the duration of clinical infectiousness (~ 3 days)), a number of secondary infections (using estimates for the reproduction number, *R*), a sampled proportion which progress to disease, a sampled proportion of infections that become hospitalised and a sampled time to hospitalisation (with different distributions for each symptom onset to hospitalisation scenario) (Figure 2, Table 2).

For the onward transmission, we explored three reproduction number values: a constant value of 0.8 or 1.2 with a range generated as +/- 5% of the constant value. For a time-varying estimate "*Rt"* we took upper/lower bounds for the 50% credible interval from a publicly available repository (16) (Supplementary 9). Mean and 95% ranges for onward transmission infections and case numbers are presented as over the 600 simulations generated from 200 simulations on each *R* value (estimate, upper and lower bound).

##### f. Reclassifying community-acquired to hospital-acquired

The number of unadjusted identified hospital-acquired COVID-19 cases is from the inflated CO-CIN dataset ("hospital-onset, hospital-acquired" cases, Figure 2, Table 1). The unadjusted community-acquired classifications were then defined as the difference between the total number of COVID-19 hospital admissions and the unadjusted identified hospital-acquired COVID-19 cases.

We adjusted the number of hospital-acquired cases by adding our model estimates of (1) "community-onset, hospital-acquired" and (2) any hospital-linked cases, to the identified hospital-acquired case numbers ("adjusted" hospital-acquired assignations). The "adjusted" community-acquired classifications are then altered accordingly. We then calculated the proportion of community cases that were reassigned as (unadjusted community # - adjusted community #) / (unadjusted community #).

To calculate the counterfactual of no transmission in hospital settings, we compared the original total number of hospitalised cases to the adjusted community number (i.e. those that we did not model as being acquired-in or linked-to hospital settings).

**Total English burden**
Acute Trusts in CO-CIN covered approximately 85% of the COVID-19 cases recorded in SUS. In order to give estimates for all English trusts, we multiplied our results by 1.17 and assumed similar levels of nosocomial transmission in non-acute English trusts.

**Supplementary 7: Symptom onset to hospitalisation**

As this was a key parameter for our estimates we chose to perform a scenario analysis around this distribution.

*Baseline scenario 1: "Best" fit to CO-CIN raw and smoothed data*

With data on 38,168 patients from CO-CIN reporting a symptom onset prior to hospitalisation in Wave 1, we could estimate the best fit to the data. However, the data suffered from "heaping" issues where patients preferably reported symptom onset data 1 week, 10 days, a fortnight or 3 weeks before hospital admission (Figure S6). This has been seen for many types of participant reported data (e.g. income (17)). To account for this we fitted to (1) the raw data (Figure S6) below using the *fitdistr R* package (18) and (2) used a penalized composite link model (19,20) to adjust for this heaping. We then compared the model fits using the Akaike Information Criterion (AIC) (21).

For both fitting to the raw and smoothed data the distribution with the smallest AIC value was the log-normal distribution (orange line in both Figure S6 and S7): AIC for the gamma distribution (next smallest AIC) was 228080 and 229646 for the smoother or raw data respectively, whilst for the log-normal distribution it was 225675 and 226842.

The values for the log-normal distribution fitted to the raw were:
Meanlog 1.662 (0.005) SDlog 0.889 (0.003)
And smoothed data:
Meanlog 1.665 (0.005) SDlog 0.894 (0.003)

We used a lognormal(1.66, 0.89) distribution in the base case Scenario 1.

*Scenario 2: previous estimates*
We also took a scenario which used a previous estimate of the time from symptom onset to hospitalisation as a gamma distribution with shape 7 and rate 1 (10) (grey line in Figure S6). This was calculated using international data from the first wave (22,23).

*Scenario 3: First Few 100 (FF100) cases in Great Britain*
We used data from the first few 100 cases data from Public Health England (11). This contains information on symptoms from the first 492 cases, 167 of which were hospitalised. At this time there was not a strict list of symptoms as there was later in 2020 (loss of taste / smell, continuous cough, fever). Fitting to this data suggested a best fit of logNormal distribution with mean log = 1.44, SD log = 0.72.

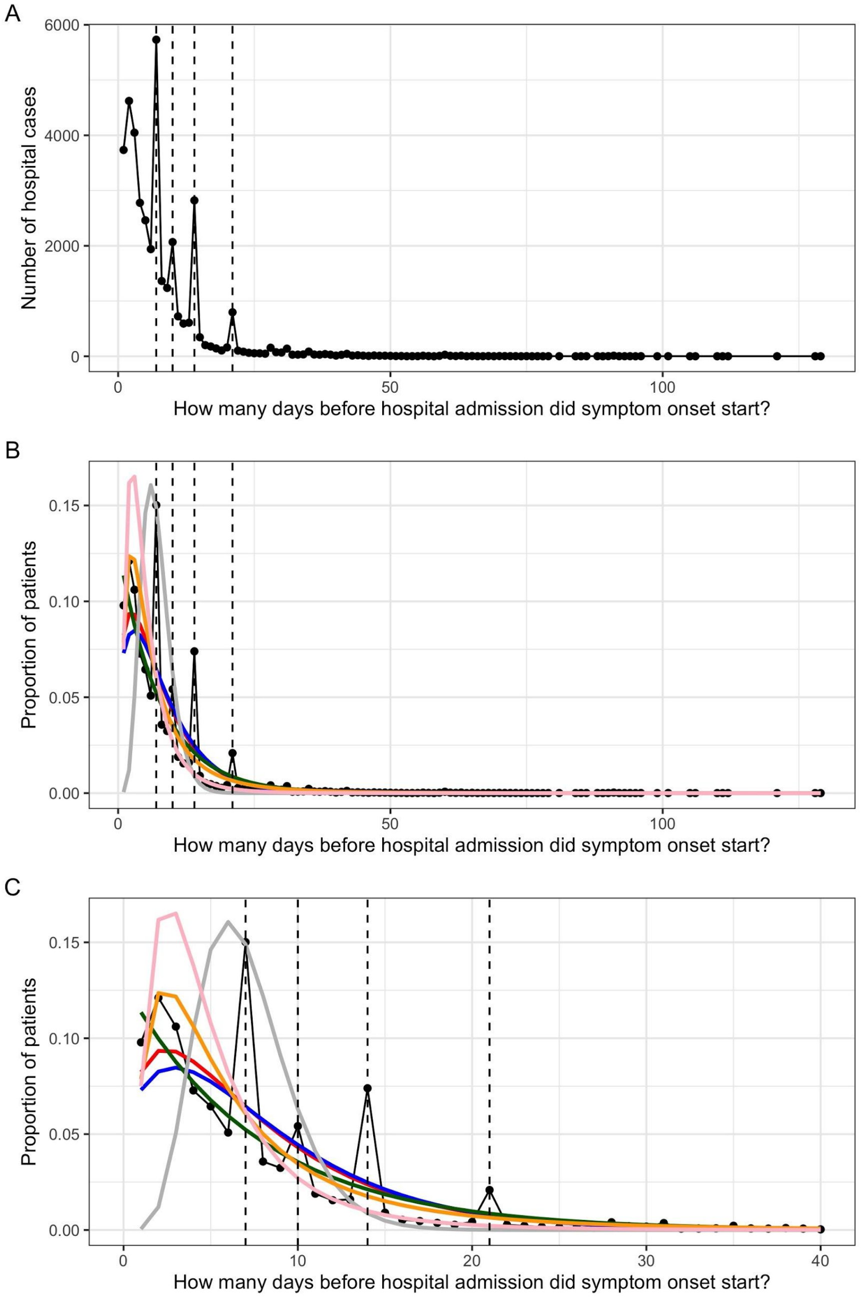


**Figure S6: What is the distribution of symptom onset before hospitalisation? (A) CO-CIN data for 38,168 patients from Wave 1 in England with a symptom onset and hospital admission date. Dashed lines indicate heaps in the data at 7, 14, 10 and 21 days prior to admission. (B) Results of probability distribution fitting to the data: red = gamma, blue = negative binomial, dark green = exponential, orange = log-normal (Scenario 1). The grey line is the distribution from** (10) **(Scenario 2: ~gamma(7,1)) and the pink line is the distribution from the FF100 data (Scenario 3: lognormal (1.44, 0.72) (C) Zoom in on (B) to show smaller differences in days between symptom onset and admission.**

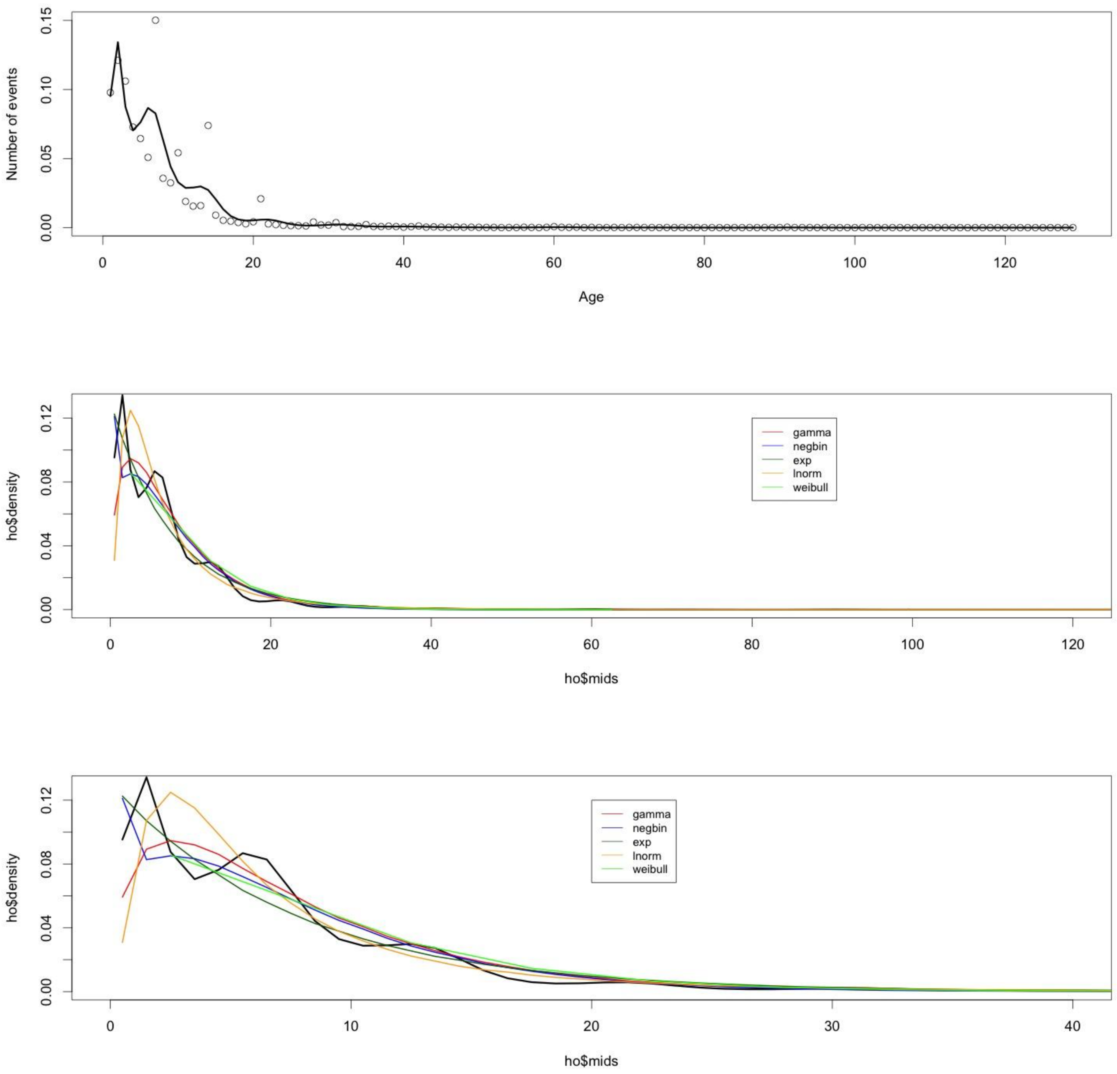


**Figure S7: What is the distribution of symptom onset before hospitalisation? (A) CO-CIN data (dots) smoothed using a penalized composite link model to give the black line. (B) Results of probability distribution fitting to the smoothed data (black line) (C) Zoom in on smaller differences between symptom onset and admission.**

### Supplementary 8:

#### Distribution of time from infection until hospital discharge for pre-symptomatic and asymptomatic patients

Let $T_{\text{inf,dis}}$ be the time from infection until discharge for pre-symptomatic and asymptomatic patients. Aim is to determine when "missed infections" will be discharged into the community. Thus, we assume that the time of infection is before discharge of the patient, i.e., $T_{\text{inf}} \leq \text{LoS}$. Given a LoS$= l$, we assume that infection is equally likely to occur on any day of length-of-stay. The distribution of $T_{\text{inf,dis}}$ is given by

$$
\begin{aligned}
P(T_{\text{inf,dis}} = t) &= P(\text{LoS} - T_{\text{inf}} = t) && (1)\\
&= \sum_{l=1}^{\infty} P(\text{LoS} - T_{\text{inf}} = t \,|\, \text{LoS} = l) p_l && (2)\\
&= \sum_{l=1}^{\infty} P(T_{\text{inf}} = l - t \,|\, \text{LoS} = l) \frac{P(\text{LoS} = l) \cdot l}{\sum_{l=1}^{\infty} l \cdot P(\text{LoS} = l)} && (3)\\
&= \sum_{l=1}^{\infty} \frac{1}{l} \mathbb{1}(t \leq l) \frac{P(\text{LoS} = l) \cdot l}{\sum_{l=1}^{\infty} l \cdot P(\text{LoS} = l)} && (4)\\
&= \sum_{l=1}^{\infty} \mathbb{1}(t \leq l) \frac{P(\text{LoS} = l)}{\sum_{l=1}^{\infty} l \cdot P(\text{LoS} = l)} && (5)\\
& && (6)
\end{aligned}
$$

where $p_l$ is the probability that on a given day, a randomly selected patient has LoS $= l$, i.e.

$$p_l = \frac{P(\text{LoS} = l) \cdot l}{\sum_{l=1}^{\infty} l \cdot P(\text{LoS} = l)}$$

**Supplementary 9: Rt estimates**

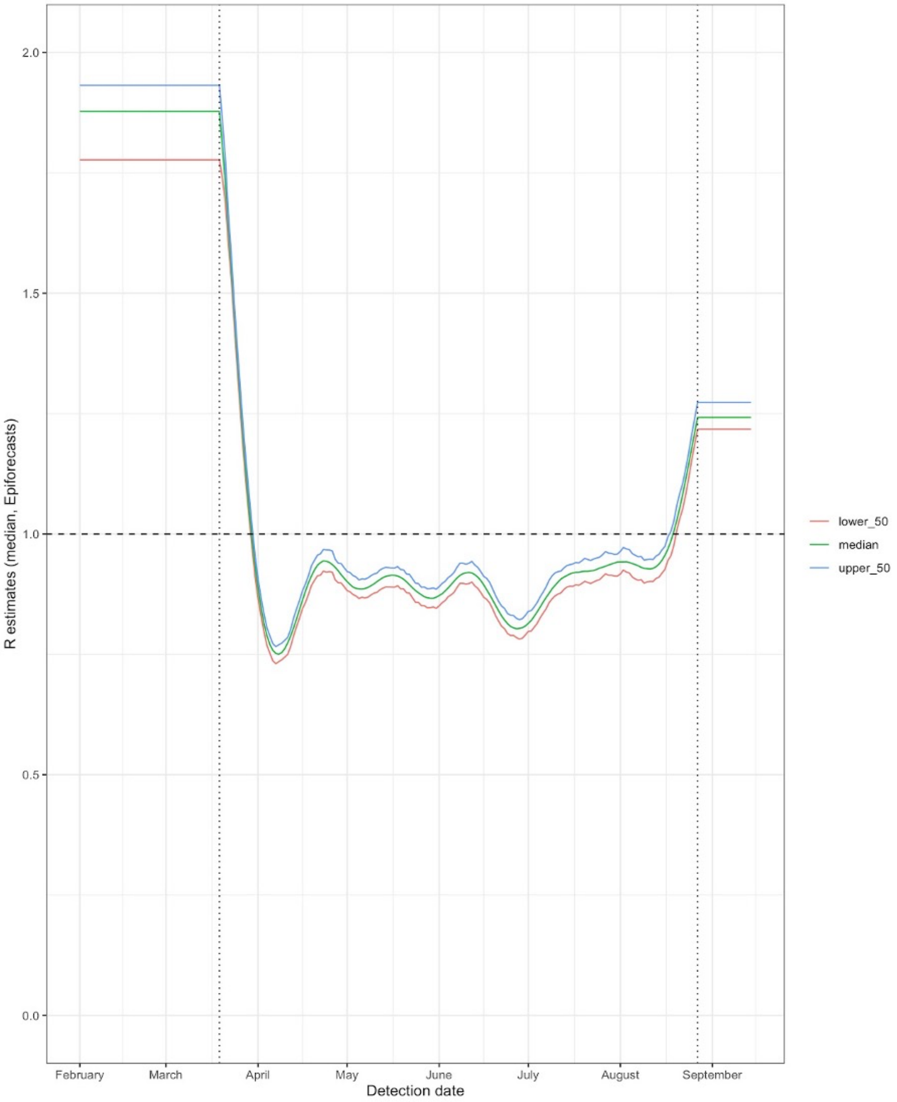


***Figure S8: Time varying estimate of Rt taken from EpiForecast team: median estimated using hospitalised cases*** [29]**(16)** ***with upper and lower bounds of the 50% credible intervals.***

Uncertainty in the simulations was generated by taking the mean and 95% ranges for onward transmission infections and case numbers are presented as over the 600 simulations generated from 200 simulations on each *R* value (estimate, upper and lower bound).

**Supplementary 10: Uncertainty inclusion**

200 simulations were generated. Each simulation included uncertainty from three stages:

Stage 1
As we generated estimates of the proportion identified by place and week, we included uncertainty from two elements each week:

(a) Length of stay distribution: bootstrap the distribution for that week from SUS. As there are so many patients (n = 237,981) in the data there is little variation produced by this variation (see Supplementary Figure S8 below, top left).
(b) Incubation period: sampled the parameters for the incubation period distribution (i.e. sample from the mean and standard deviation for the lognormal distribution from a normal distribution with the estimated mean and sd to give a different distribution for each sample for the time to symptom onset from infection, see Table 2).

This incubation period distribution and length of stay for non-COVID patients was used for the entire of the simulation. This is coded in “trust_proportion_detect_by_week_all.R” (4). It gives the variation in the proportion of hospital-acquired infections identified and is presented in Figure 3c, and shown again in Figure S9 for a cutoff of symptom onset more than 7 days from admission.

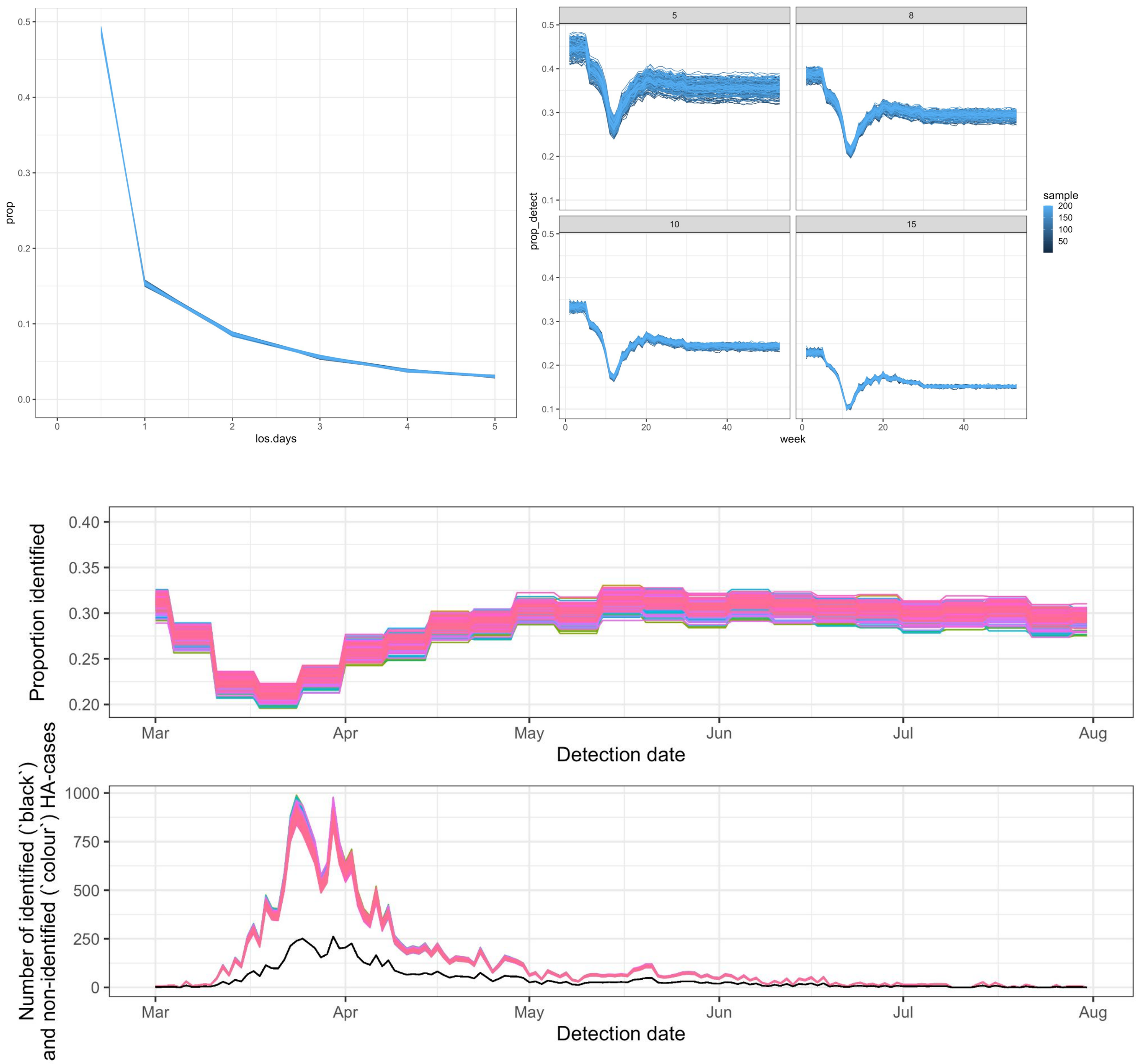


**Figure S9:** Uncertainty in the length of stay (top left) and incubation period drive uncertainty in the proportion identified (top right and middle). The inverse of this proportion multiplies the number of identified hospital-acquired cases per week (black, bottom) to calculate the number of unidentified infections (colour, bottom).

For example, towards the end of March: 250 hospital-acquired cases were identified in the inflated CO-CIN (Figure S9, bottom). At this stage it is likely that we were identifying between 20% and 22% of hospital-acquired cases (Figure S9, top). Hence this corresponds to between 840 and 1,000 missed cases.

Stage 2

To accounting for binomial sampling variation, the proportion identified for each sample and week (generated above) were used within a Bayesian framework as the binomial probability of identification to infer from the number of identified hospital-acquired cases, the total number of hospital-acquired infections ("trials").

In more detail - using the distributions in step 1 within our function we could generate 200 samples of the proportion of true hospital acquired infections that were identified each week, *i*, and setting, *j*, from hospital data ($p_{i,j}$). Assuming the number of hospital acquired infections were binomially distributed, we estimated the weekly number of true hospital-acquired infections, $X_{i,j} \sim \text{Bin}(N_{i,j}, p_{i,j})$. Subtracting from this the identified weekly hospital-acquired infection numbers we can estimate the number of unidentified hospital-acquired infections.

This used the function in "binom_posterior.R" (4)**.**

Stage 3

The uncertainty in the natural history trajectory for each of these unidentified hospital-acquired infections was then calculated (as shown in Figure 2d) by sampling from the relevant distributions for the probability (e.g. of returning as a hospitalised cases) and timings (e.g. symptom onset after infection). This is coded in "perc_contribution_function_trust_week.R" (4).

For each unidentified infection, the probability of returning as a COVID-19 case to hospital is a Bernoulli trial for each missed infection with weekly randomly sampled probability of returning taken from a uniform distribution over 10-15%. This probability of a "missed" unidentified infection returning of a community infection becoming hospitalised is fixed across each of the 200 simulations.

Each of the following timings for each returning to hospital as a case unidentified hospital-acquired infection are then sampled from the relevant distributions (Table 2):

(a) From infection to discharge
(b) From infection to symptom onset (incubation period)
(c) From symptom onset to hospitalisation (this is scenario dependent)

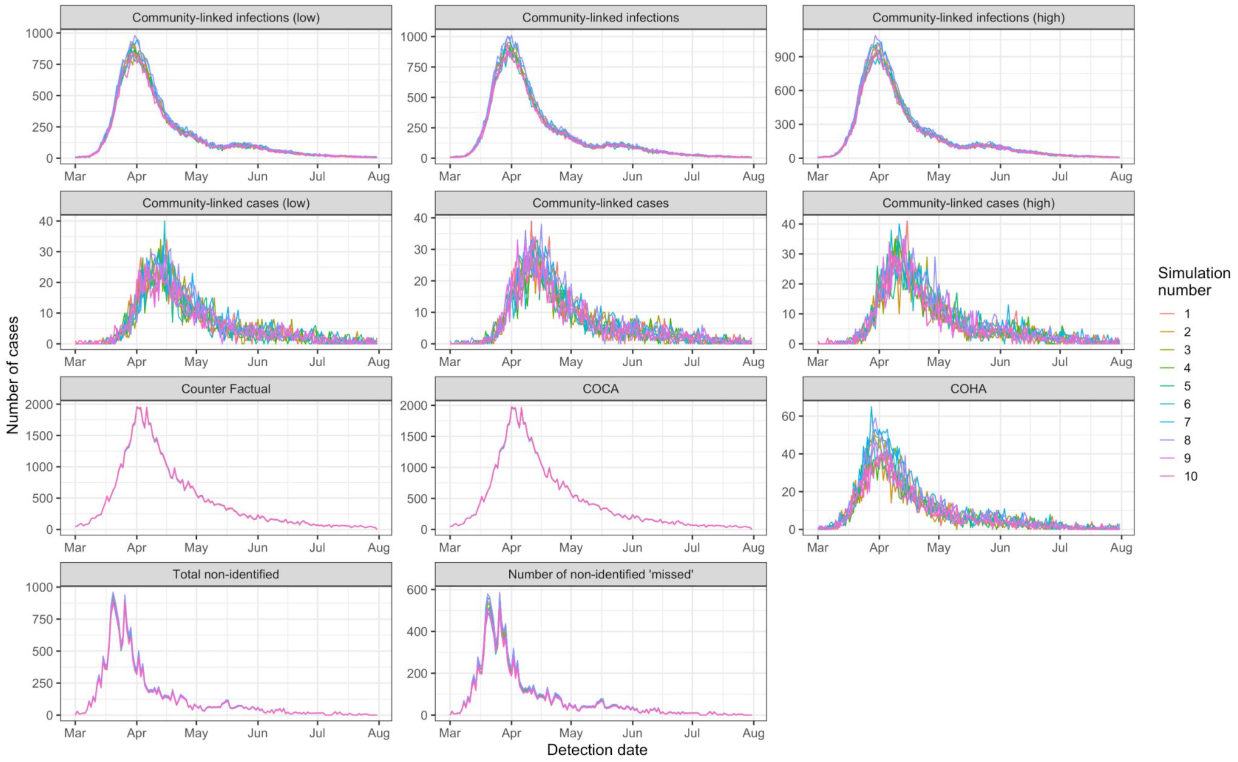


**Figure S10:** Example time series with a cutoff of at least 7 days from symptom onset to hospitalisation for defining a hospital-acquired case: the first 10 simulations for key model outputs are shown in the 10 colours in the above facets over time (detection date). The top two rows show the variation in community-onset, hospital-linked infections (first row) and subsequent cases (second row) at low, mean and high values of onward transmission ($R$ = 0.76, 0.8, 0.84). The third row shows the counterfactual: the number of hospitalised cases there would be predicted to be without any hospital-acquisition of SARS-CoV-2, alongside the community-onset, community-acquired ("COCA") and community-onset, hospital-acquired ("COHA) case estimates. The final row shows the same variation shown in Figure S8: the total number of unidentified infections and the "missed" subset of these ("missed" due to discharge prior to symptom onset).

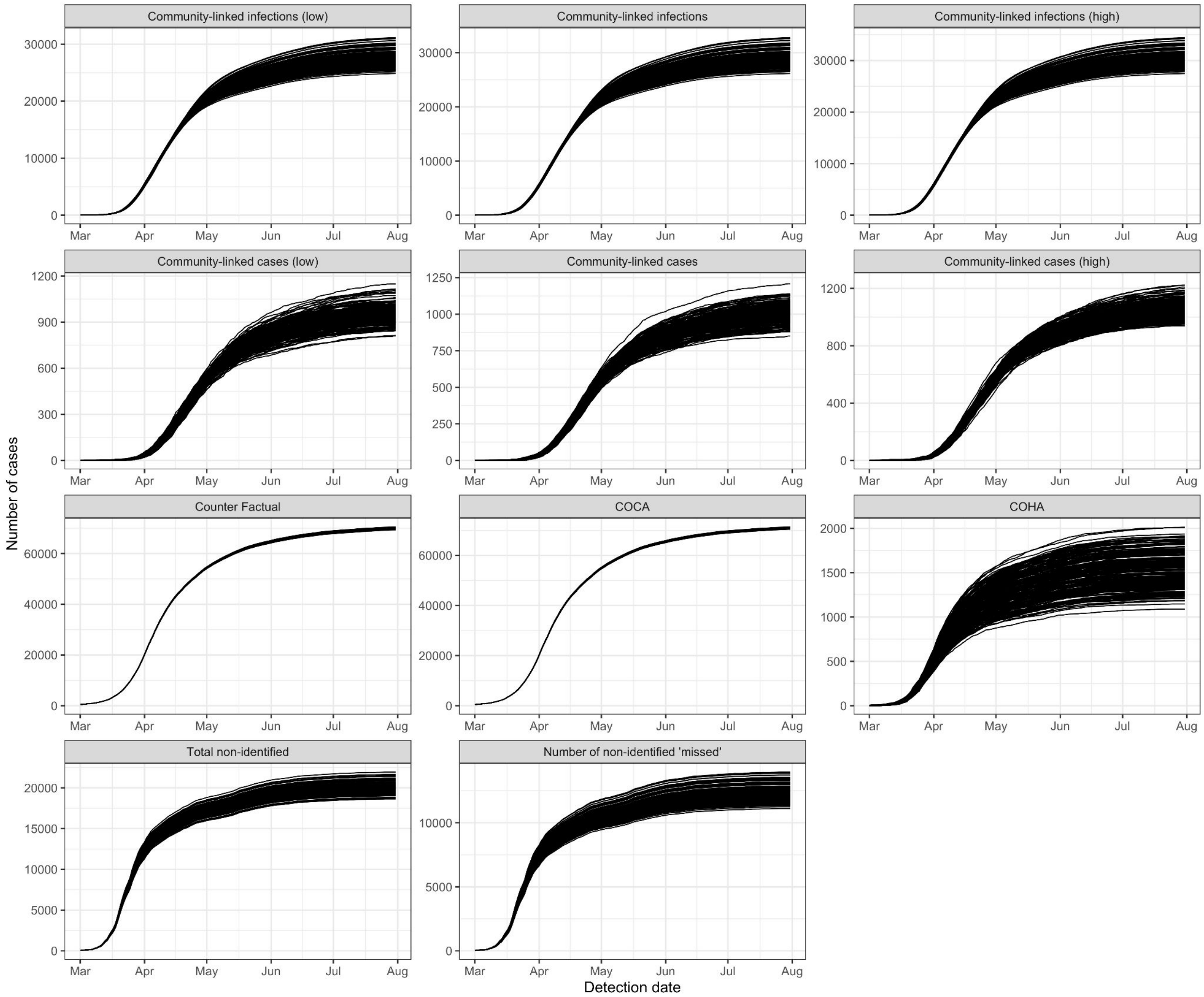


**Figure S11:** Example cumulative values as in Figure S10 for all 200 simulations (each black line) with a cutoff of at least 7 days from symptom onset to hospitalisation for defining a hospital-acquired case. The top two rows show the variation in community-onset, hospital-linked infections (first row) and subsequent cases (second row) at low, mean and high values of onward transmission ($R$ = 0.76, 0.8, 0.84). The third row shows the counterfactual: the number of hospitalised cases there would be predicted to be without any hospital-acquisition of SARS-CoV-2, alongside the community-onset, community-acquired ("COCA") and community-onset, hospital-acquired ("COHA) case estimates. The final row shows the same variation shown in Figure S8: the total number of unidentified infections and the "missed" subset of these ("missed" due to discharge prior to symptom onset). Note the variation in the y axis values.

We decided to use 200 simulations as above approximately 150 simulations the output for key parameters (shown in Figure S12) stabilised.

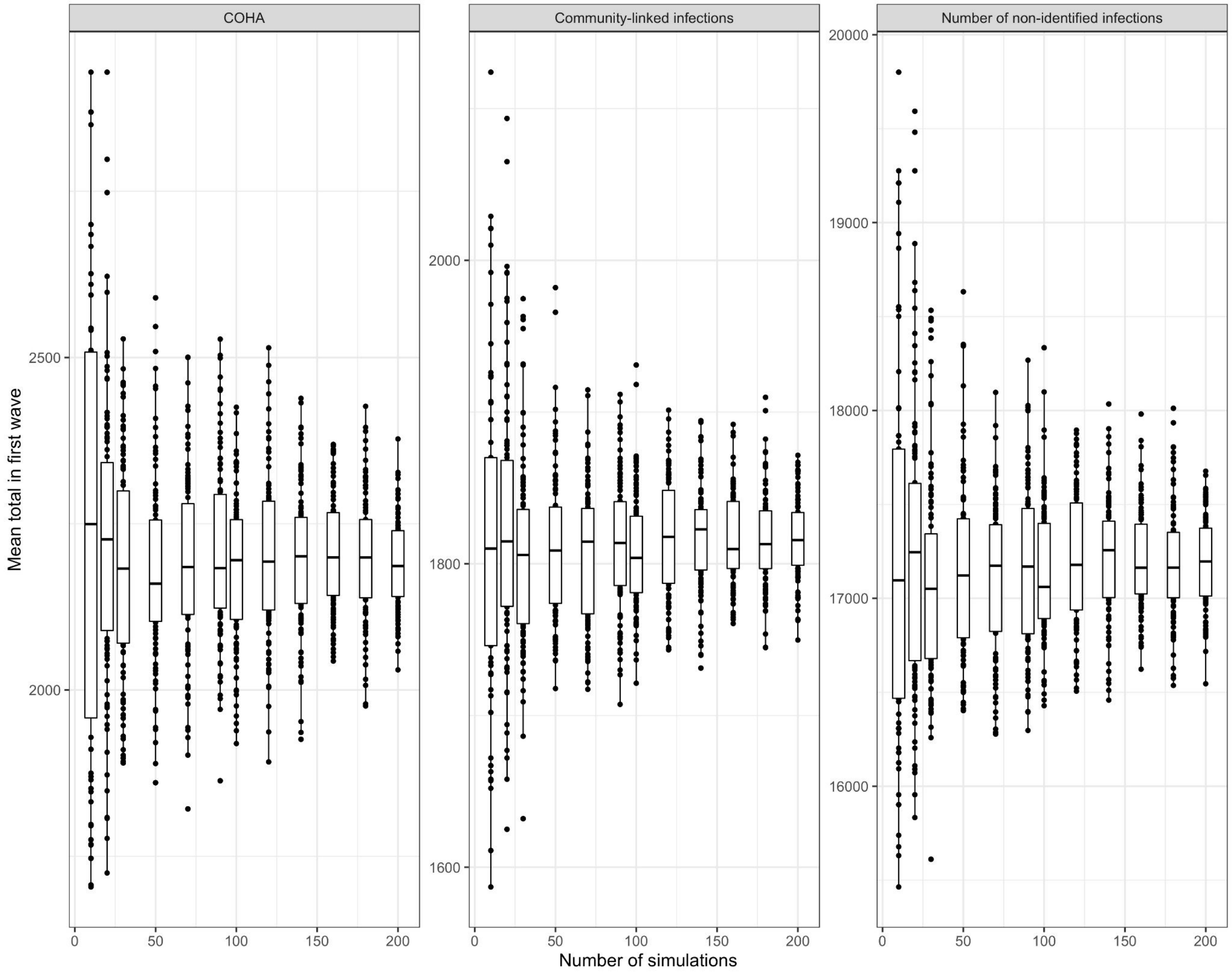


**Figure S12:** Boxplot of mean total value of key outcome variables over the "first wave" (to 31st July 2020) against the number of simulations. Left is "community onset, hospital-acquired" cases (COHA), middle are community-linked infections and right is the number of unidentified infections.

**Conclusion**

Uncertainty in our estimates was generated from sampling from a range of natural history distributions and the length of stay data. As we had data from SUS on the latter for a large number of non-COVID patients, we had little ambiguity in this key parameter for estimating the proportion of hospital-acquired infections identified. Moreover, much of the uncertainty was in the timing of events (symptom onset 2 or 5 days from infection for example), which, when aggregated over a 7-month period had little impact on the final aggregated results.

**Supplementary 11: Additional results**

- **Figure 5 additional analysis**
- **Table S3**: Additional reported results
- **Table S4**: Estimated percentage of “community onset, community acquired” infections that would be re-classified as “community onset, hospital acquired” infections
- **Table S5**: Estimated number of community onset, hospital-linked cases
- **Figure S13:** Impact of 1 vs 5 day discharge before associated identified hospital case
- **Figure S14:** Impact of R value variation over time (not just aggregated)

**Figure 5 additional analysis**

- Of all hospital patients who had a SARS-CoV-2 infection some time during their stay, 29.6% (28.9%, 30.5%) were hospital-acquired (E/(A+D), Figure 5).
- With the addition of hospital-linked infections, out of all hospital patients with a SARS-CoV-2 infection, 31.5% (30.6%, 32.4%) were estimated to have acquired their infection in hospitals or were hospital-linked ((E+F)/(A+D), Figure 5).

**Table S3:** Estimated additional main results for 14 and 4 day cut-offs in line with 7 day values in main text

| Estimate | Cutoff<br>7 | <br>14 | <br>4 | <br>Details |
|---|---|---|---|---|
| "hospital-onset, hospital-acquired" identified cases across acute English Trusts up to the 31st July 2020 | 6,640 | 4,440 | 7,830 | From adjusted CO-CIN |
| unidentified hospital-acquired infections | 20,000 (19,200, 21,100) | 29,000 (28,400, 29,600) | 17,500 (16,000, 19,300) | mean; 95% range over 200 simulations |
| Percentage of "community-onset, community-acquired" that should be classified as "community-onset, hospital-acquired" | 2.1% (1.7%, 2.6%) | 2.6% (2.1%, 3.1%) | 2.1% (1.7%, 2.6%) | mean; 95% range over 200 simulations: |
| "community-onset, hospital-linked" cases | 1,600 (1,600, 1,700) | 2,100 (2,000, 2,200) | 1,600 (1,400, 1,700) | For the time varying $R$ value mean; 95% range over 600 simulations |

**Table S4:** Estimated mean and 95% quantile range over 200 simulations of the percentage of "community onset, community acquired" infections that would be re-classified as "community onset, hospital acquired" infections under different *R* values, hospital-acquired (HA) definition cutoffs (if symptom onset starts this many days from admission), discharge times from associated hospital-acquired case for unidentified hospital-acquired infection and scenarios for symptom onset to hospitalisation.

| R value (0.8, 1.2, *rt*) | HA definition | Discharge time for | Symptom onset to hospitalisation scenario | Mean | 95% quantile range | |
|---|---|---|---|---|---|---|
| 0.8 | 5 | 5 | 1 | 2.1 | 1.6 | 2.5 |
| 0.8 | 5 | 5 | 2 | 2.1 | 1.6 | 2.5 |
| 0.8 | 5 | 5 | 3 | 2.1 | 1.6 | 2.5 |
| 0.8 | 8 | 5 | 1 | 2.1 | 1.7 | 2.6 |
| 0.8 | 8 | 5 | 2 | 2.1 | 1.7 | 2.6 |
| 0.8 | 8 | 5 | 3 | 2.1 | 1.7 | 2.6 |
| 0.8 | 15 | 5 | 1 | 2.6 | 2.1 | 3.1 |
| 0.8 | 15 | 5 | 2 | 2.6 | 2.1 | 3.1 |
| 0.8 | 15 | 5 | 3 | 2.6 | 2.1 | 3.1 |
| 1.2 | 5 | 5 | 1 | 2.1 | 1.6 | 2.5 |
| 1.2 | 5 | 5 | 2 | 2.1 | 1.6 | 2.5 |
| 1.2 | 5 | 5 | 3 | 2.1 | 1.6 | 2.5 |
| 1.2 | 8 | 5 | 1 | 2.1 | 1.7 | 2.6 |
| 1.2 | 8 | 5 | 2 | 2.1 | 1.7 | 2.6 |
| 1.2 | 8 | 5 | 3 | 2.1 | 1.7 | 2.6 |
| 1.2 | 15 | 5 | 1 | 2.6 | 2.1 | 3.1 |
| 1.2 | 15 | 5 | 2 | 2.6 | 2.1 | 3.1 |
| 1.2 | 15 | 5 | 3 | 2.6 | 2.1 | 3.1 |
| rt | 5 | 5 | 1 | 2.1 | 1.6 | 2.6 |
| rt | 5 | 5 | 2 | 2.1 | 1.6 | 2.5 |
| rt | 5 | 5 | 3 | 2.1 | 1.6 | 2.6 |
| rt | 8 | 5 | 1 | 2.1 | 1.7 | 2.6 |
| rt | 8 | 5 | 2 | 2.1 | 1.7 | 2.6 |
| rt | 8 | 5 | 3 | 2.1 | 1.7 | 2.6 |

| | | | | | | |
|---|---|---|---|---|---|---|
| rt | 15 | 5 | 1 | 2.6 | 2.1 | 3.1 |
| rt | 15 | 5 | 2 | 2.6 | 2.1 | 3.1 |
| rt | 15 | 5 | 3 | 2.6 | 2.1 | 3.1 |
| 0.8 | 5 | 1 | 1 | 2.1 | 1.6 | 2.5 |
| 0.8 | 5 | 1 | 2 | 2.1 | 1.6 | 2.5 |
| 0.8 | 5 | 1 | 3 | 2.1 | 1.6 | 2.5 |
| 0.8 | 8 | 1 | 1 | 2.1 | 1.7 | 2.6 |
| 0.8 | 8 | 1 | 2 | 2.1 | 1.7 | 2.6 |
| 0.8 | 8 | 1 | 3 | 2.1 | 1.7 | 2.6 |
| 0.8 | 15 | 1 | 1 | 2.6 | 2.1 | 3.1 |
| 0.8 | 15 | 1 | 2 | 2.6 | 2.1 | 3.1 |
| 0.8 | 15 | 1 | 3 | 2.6 | 2.1 | 3.1 |
| 1.2 | 5 | 1 | 1 | 2.1 | 1.6 | 2.6 |
| 1.2 | 5 | 1 | 2 | 2.1 | 1.6 | 2.5 |
| 1.2 | 5 | 1 | 3 | 2.1 | 1.6 | 2.5 |
| 1.2 | 8 | 1 | 1 | 2.1 | 1.7 | 2.6 |
| 1.2 | 8 | 1 | 2 | 2.1 | 1.7 | 2.6 |
| 1.2 | 8 | 1 | 3 | 2.1 | 1.7 | 2.6 |
| 1.2 | 15 | 1 | 1 | 2.6 | 2.1 | 3.1 |
| 1.2 | 15 | 1 | 2 | 2.6 | 2.1 | 3.1 |
| 1.2 | 15 | 1 | 3 | 2.6 | 2.1 | 3.1 |
| rt | 5 | 1 | 1 | 2.1 | 1.6 | 2.5 |
| rt | 5 | 1 | 2 | 2.1 | 1.6 | 2.5 |
| rt | 5 | 1 | 3 | 2.1 | 1.6 | 2.5 |
| rt | 8 | 1 | 1 | 2.1 | 1.7 | 2.6 |
| rt | 8 | 1 | 2 | 2.1 | 1.7 | 2.6 |
| rt | 8 | 1 | 3 | 2.1 | 1.7 | 2.6 |
| rt | 15 | 1 | 1 | 2.6 | 2.1 | 3.1 |

| | | | | | | |
|---|---|---|---|---|---|---|
| rt | 15 | 1 | 2 | 2.6 | 2.1 | 3.1 |
| rt | 15 | 1 | 3 | 2.6 | 2.1 | 3.1 |

**Table S5:** Estimated mean and 95% quantile range over 200 simulations number and percentage contribution of "community onset, hospital linked cases" under different *R* values, hospital-acquired (HA) definition cutoffs (if symptom onset starts this many days from admission), discharge times from associated hospital-acquired case for unidentified hospital-acquired infection and scenarios for symptom onset to hospitalisation.

| | | | | **Number of infections** | | | **Proportion of community onset community acquired cases** | | |
|---|---|---|---|---|---|---|---|---|---|
| **R value (0.8, 1.2, *rt*)** | **HA definition cutoff (5,8,15)** | **Discharge time for unidentified hospital-acquired infection (1 or 5)** | **Symptom onset to hospitalisation scenario (1-3)** | **Mean** | **95% quantile range** | | **Mean** | **95% quantile range** | |
| 0.8 | 5 | 5 | 1 | 1000 | 800 | 1100 | 1.3 | 1.2 | 1.5 |
| 0.8 | 5 | 5 | 2 | 1000 | 800 | 1100 | 1.3 | 1.2 | 1.6 |
| 0.8 | 5 | 5 | 3 | 1000 | 800 | 1100 | 1.3 | 1.2 | 1.5 |
| 0.8 | 8 | 5 | 1 | 1000 | 900 | 1100 | 1.4 | 1.2 | 1.5 |
| 0.8 | 8 | 5 | 2 | 1000 | 900 | 1100 | 1.4 | 1.3 | 1.5 |
| 0.8 | 8 | 5 | 3 | 1000 | 900 | 1100 | 1.4 | 1.2 | 1.5 |
| 0.8 | 15 | 5 | 1 | 1300 | 1200 | 1400 | 1.7 | 1.6 | 1.8 |
| 0.8 | 15 | 5 | 2 | 1300 | 1200 | 1400 | 1.7 | 1.6 | 1.8 |
| 0.8 | 15 | 5 | 3 | 1300 | 1200 | 1400 | 1.7 | 1.6 | 1.8 |
| 1.2 | 5 | 5 | 1 | 2600 | 2300 | 2900 | 3.6 | 3.2 | 4.1 |
| 1.2 | 5 | 5 | 2 | 2600 | 2300 | 2900 | 3.6 | 3.3 | 4.1 |
| 1.2 | 5 | 5 | 3 | 2600 | 2300 | 3000 | 3.7 | 3.3 | 4.2 |
| 1.2 | 8 | 5 | 1 | 2700 | 2500 | 3000 | 3.8 | 3.5 | 4.1 |
| 1.2 | 8 | 5 | 2 | 2700 | 2500 | 3000 | 3.8 | 3.5 | 4.1 |
| 1.2 | 8 | 5 | 3 | 2700 | 2500 | 3000 | 3.8 | 3.5 | 4.1 |
| 1.2 | 15 | 5 | 1 | 3400 | 3200 | 3700 | 4.6 | 4.3 | 4.9 |
| 1.2 | 15 | 5 | 2 | 3400 | 3300 | 3700 | 4.6 | 4.4 | 4.9 |

| | | | | | | | | | |
|---|---|---|---|---|---|---|---|---|---|
| 1.2 | 15 | 5 | 3 | 3500 | 3200 | 3700 | 4.6 | 4.3 | 5 |
| rt | 5 | 5 | 1 | 1600 | 1400 | 1700 | 2.2 | 2 | 2.4 |
| rt | 5 | 5 | 2 | 1600 | 1400 | 1700 | 2.2 | 2 | 2.4 |
| rt | 5 | 5 | 3 | 1600 | 1400 | 1700 | 2.2 | 2 | 2.4 |
| rt | 8 | 5 | 1 | 1600 | 1600 | 1700 | 2.3 | 2.1 | 2.4 |
| rt | 8 | 5 | 2 | 1600 | 1500 | 1700 | 2.3 | 2.1 | 2.4 |
| rt | 8 | 5 | 3 | 1600 | 1500 | 1800 | 2.3 | 2.1 | 2.4 |
| rt | 15 | 5 | 1 | 2100 | 2000 | 2200 | 2.8 | 2.7 | 2.9 |
| rt | 15 | 5 | 2 | 2100 | 2000 | 2200 | 2.8 | 2.7 | 2.9 |
| rt | 15 | 5 | 3 | 2100 | 2000 | 2200 | 2.8 | 2.7 | 2.9 |
| 0.8 | 5 | 1 | 1 | 900 | 800 | 1100 | 1.3 | 1.2 | 1.5 |
| 0.8 | 5 | 1 | 2 | 1000 | 800 | 1100 | 1.3 | 1.2 | 1.5 |
| 0.8 | 5 | 1 | 3 | 1000 | 800 | 1100 | 1.3 | 1.2 | 1.5 |
| 0.8 | 8 | 1 | 1 | 1000 | 900 | 1100 | 1.4 | 1.3 | 1.5 |
| 0.8 | 8 | 1 | 2 | 1000 | 900 | 1100 | 1.4 | 1.3 | 1.5 |
| 0.8 | 8 | 1 | 3 | 1000 | 900 | 1100 | 1.4 | 1.3 | 1.5 |
| 0.8 | 15 | 1 | 1 | 1300 | 1200 | 1400 | 1.7 | 1.6 | 1.8 |
| 0.8 | 15 | 1 | 2 | 1300 | 1200 | 1400 | 1.7 | 1.6 | 1.8 |
| 0.8 | 15 | 1 | 3 | 1300 | 1200 | 1400 | 1.7 | 1.6 | 1.8 |
| 1.2 | 5 | 1 | 1 | 2600 | 2300 | 2900 | 3.6 | 3.2 | 4.1 |
| 1.2 | 5 | 1 | 2 | 2600 | 2300 | 2900 | 3.6 | 3.2 | 4.1 |

| | | | | | | | | | |
|---|---|---|---|---|---|---|---|---|---|
| 1.2 | 5 | 1 | 3 | 2600 | 2300 | 2900 | 3.6 | 3.3 | 4.1 |
| 1.2 | 8 | 1 | 1 | 2700 | 2500 | 2900 | 3.8 | 3.5 | 4.1 |
| 1.2 | 8 | 1 | 2 | 2700 | 2500 | 3000 | 3.8 | 3.5 | 4.1 |
| 1.2 | 8 | 1 | 3 | 2700 | 2500 | 3000 | 3.8 | 3.5 | 4.1 |
| 1.2 | 15 | 1 | 1 | 3400 | 3200 | 3700 | 4.6 | 4.3 | 4.9 |
| 1.2 | 15 | 1 | 2 | 3400 | 3200 | 3700 | 4.6 | 4.3 | 4.9 |
| 1.2 | 15 | 1 | 3 | 3400 | 3200 | 3700 | 4.6 | 4.3 | 4.9 |
| *rt* | 5 | 1 | 1 | 1300 | 1200 | 1500 | 1.9 | 1.7 | 2.1 |
| *rt* | 5 | 1 | 2 | 1300 | 1200 | 1500 | 1.9 | 1.7 | 2.1 |
| *rt* | 5 | 1 | 3 | 1300 | 1200 | 1500 | 1.9 | 1.7 | 2.1 |
| *rt* | 8 | 1 | 1 | 1400 | 1300 | 1500 | 1.9 | 1.8 | 2.1 |
| *rt* | 8 | 1 | 2 | 1400 | 1300 | 1500 | 1.9 | 1.8 | 2.1 |
| *rt* | 8 | 1 | 3 | 1400 | 1300 | 1500 | 1.9 | 1.8 | 2.1 |
| *rt* | 15 | 1 | 1 | 1800 | 1700 | 1900 | 2.4 | 2.2 | 2.5 |
| *rt* | 15 | 1 | 2 | 1800 | 1700 | 1900 | 2.4 | 2.2 | 2.5 |
| *rt* | 15 | 1 | 3 | 1800 | 1700 | 1900 | 2.4 | 2.2 | 2.5 |

#### Impact of 1 - 5 day discharge

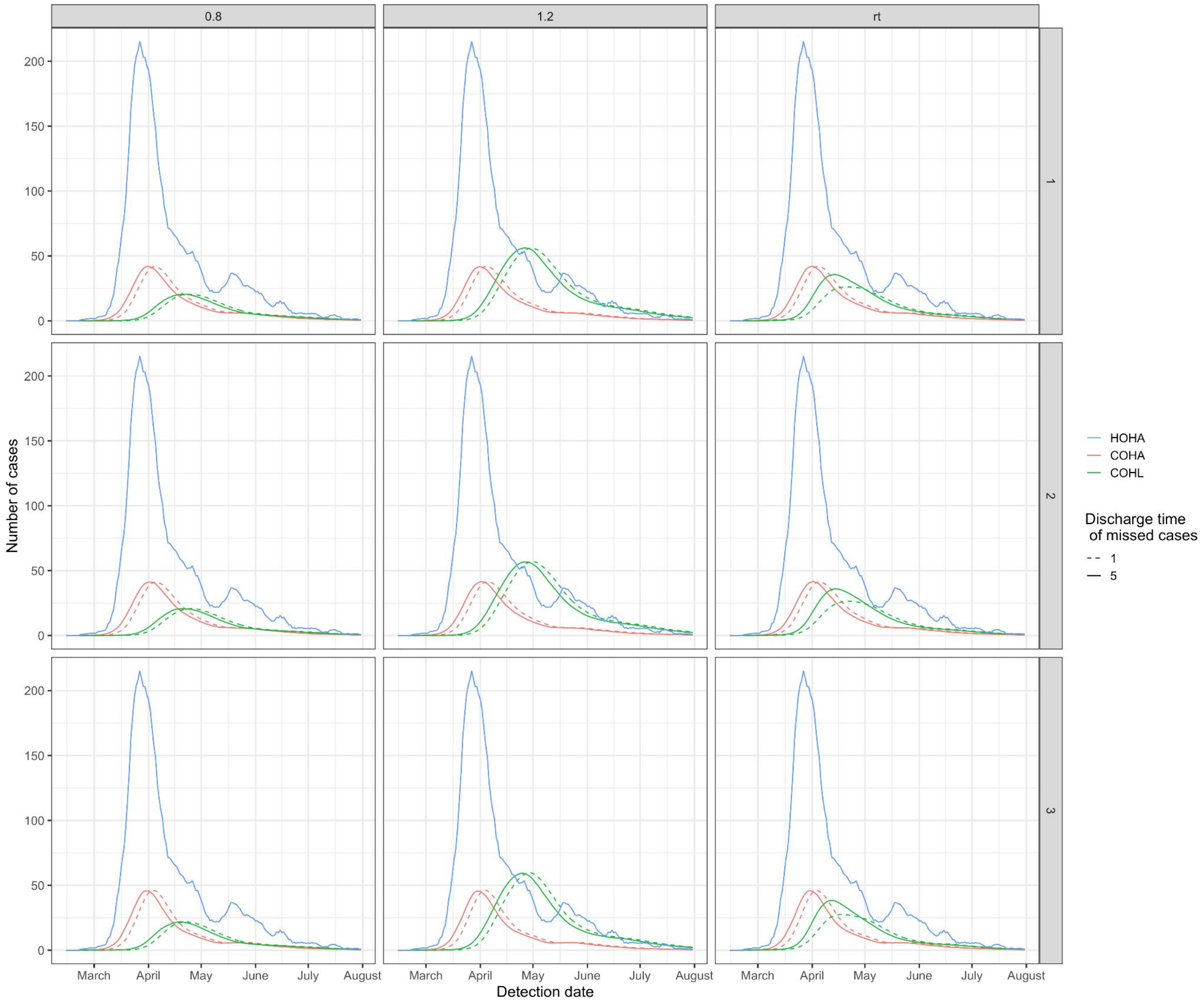


**Figure S13: The impact of discharging missed cases 5 days (solid line, baseline) or 1 day (dashed line) before the associated identified hospital-acquired case at a cut-off threshold of 7 days from admission across different *R* values (columns) and Scenarios (rows) of symptom onset to hospitalisation. This is for "hospital-onset, hospital-acquired" (HOHA, blue), "community-onset, hospital-acquired" (COHA, red) and "community-onset, hospital-linked" (COHL, green) cases**

As shown in Figure S13, there is a minimal impact of varying the day of discharge of missed cases, except for the "community-onset, hospital-linked" (COHL) cases when using the time varying *R* estimates ("rt"). Cumulatively, up to the end of July 2020, this results in a less than 0.001% change in the number of "community-onset, community-acquired" cases but a ~30% higher number of "community-onset, hospital-linked" cases when using the time varying *R* estimates ("rt") and a 5 day discharge. This is due to a synergistic impact of the missed infections entering the community at peak R value (before early April).

#### Impact of R value variation

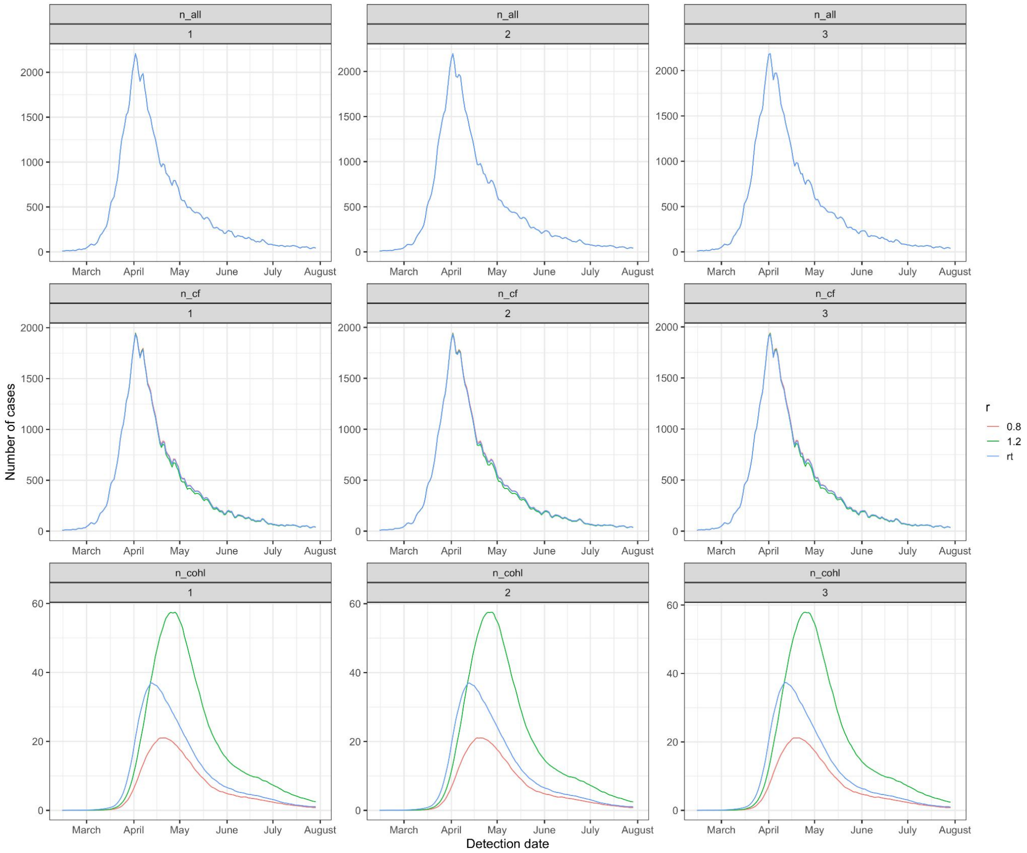


**Figure S14: Time series of all hospitalised, counter-factual and community onset, hospital-linked cases under different onward transmission (*R*) values (median values shown here, colours). This is for a cutoff of 7 days from admission for the hospital-acquired definition and for the three scenarios (columns) for symptom onset to hospitalisation.**

**Supplementary analysis 12: Grouped Trust level analysis**

**Method:** We applied the same analysis (shown in Figure 2) at the individual acute Trust level (n = 126) and then aggregated the results. We performed this analysis for all three cutoffs, three *R* values and for the first symptom onset to hospitalisation scenario with 50 simulations for each of the 126 Trusts to generate uncertainty ranges.

**Results:**
4 Trusts had no nosocomial cases recorded in the data over this time period. Two Trusts had no nosocomial cases recorded when using a 14 day cutoff for definition of a nosocomial case.

The proportion missed each week varied over trusts with a mean of 29% and a range between 0 and 88% over 50 simulations and all weeks and Trusts. 0.2% of the proportion detected estimates were zero.

Comparing the aggregated England setting (data pooled before doing analysis) to the grouped individual Trust (analysis performed at the Trust level and then aggregated) analysis shows similar results but the levels from the individual Trust analysis is higher (Table S5). Some variation would be expected due to rounding e.g. of the number of missed infections from the identified number of hospital-acquired cases. At the baseline cutoff of symptom onset 8 or more days from admission, the variation is relatively small, but it increases at a 15 or more days from admission cutoff, especially for "community-onset, hospital-linked" cases. The similarity in key indicators is shown in Figure S15-17 below. Using the grouped individual Trust analysis predicts that 25.5% (24.6%, 26.4%) of identified COVID-19 cases in hospitals were hospital-acquired, higher than the level predicted from the aggregated England setting: 20.1% (19.2%, 20.7%).

**Comparison and interpretation:** The proportion identified is predicted to be very small when there are few hospital-onset, hospital-acquired (HOHA) cases, as is often the case when doing the analysis at the individual trust level. Using the Bayesian framework to infer the total number of hospital-acquired infections ("trials") results in higher numbers (~50%) for the estimated number of unidentified hospital-acquired infections and hence onward case estimates (COHL / COHA). For example, 1 HOHA case, with a proportion detected of 0.005, is predicted to be linked to 524 hospital-acquired infections. However, 7 HOHA cases, with the same proportion detected, results in a predicted 1450 infections: an increase of 3x infections instead of 7x as might be expected from the increase in HOHA. We believe that the analysis at the Trust level suffers from issues of small numbers and issues with using the empiric length of stay distributions. This leads to unrealistically small proportions detected and hence inflation to a greater number of missed infections.

***Table S5: Comparison of England level vs. grouped trusts analysis for varying cutoff and R values for three key indicators under the first scenario for symptom onset to hospital admission. The values presented are the mean and 95% quantile over 200 simulations for the aggregated England setting and over 50 simulations for each Trust. Bold values are those with the baseline cutoff.***

| Setting | Cutoff for defining hospital-acquired (symptom onset on this day or later after admission) | *R* value | Number of hospital-onset hospital-acquired identified cases (HOHA) | Number of unidentified hospital-acquired infections | Number of community-onset hospital-linked cases (COHL) |
|---|---|---|---|---|---|
| ENG | 5 | 0.8 | 7,800 (7,800, 7,800) | 17,400 (16,100, 19,100) | 1,000 (900, 1,100) |
| grouped | | 0.8 | 7,400 (7,400, 7,400) | 24,700 (22,500, 27,100) | 1,500 (1,400, 1,700) |
| ENG | | 1.2 | 7,800 (7,800, 7,800) | 17,400 (16,100, 19,100) | 2,600 (2,400, 2,900) |
| grouped | | 1.2 | 7,400 (7,400, 7,400) | 24,700 (22,500, 27,100) | 3,400 (3,100, 3,900) |
| ENG | | rt | 7,800 (7,800, 7,800) | 17,400 (16,100, 19,100) | 1,600 (1,500, 1,700) |
| grouped | | rt | 7,400 (7,400, 7,400) | 24,700 (22,500, 27,100) | 2,400 (2,200, 2,600) |
| **ENG** | **8** | **0.8** | **6,600 (6,600, 6,600)** | **20,000 (19,300, 21,000)** | **1,000 (900, 1,100)** |
| **grouped** | | **0.8** | **6,200 (6,200, 6,200)** | **29,200 (28,100, 30,800)** | **1,700 (1,500, 1,800)** |
| **ENG** | | **1.2** | **6,600 (6,600, 6,600)** | **20,000 (19,300, 21,000)** | **2,800 (2,500, 3,100)** |
| **grouped** | | **1.2** | **6,200 (6,200, 6,200)** | **29,200 (28,100, 30,800)** | **3,800 (3,500, 4,200)** |
| **ENG** | | **rt** | **6,600 (6,600, 6,600)** | **20,000 (19,300, 21,000)** | **1,700 (1,600, 1,800)** |
| **grouped** | | **rt** | **6,200 (6,200, 6,200)** | **29,200 (28,100, 30,800)** | **2,500 (2,400, 2,700)** |
| ENG | 15 | 0.8 | 4,400 (4,400, 4,400) | 29,100 (28,500, 29,700) | 1,300 (1,200, 1,400) |
| grouped | | 0.8 | 4,000 (4,000, 4,000) | 55,700 (51,500, 62,900) | 2,600 (2,400, 3,000) |
| ENG | | 1.2 | 4,400 (4,400, 4,400) | 29,100 (28,500, 29,700) | 3,500 (3,300, 3,800) |
| grouped | | 1.2 | 4,000 (4,000, 4,000) | 55,700 (51,500, 62,900) | 6,600 (5,900, 7,400) |
| ENG | | rt | 4,400 (4,400, 4,400) | 29,100 (28,500, 29,700) | 2,100 (2,000, 2,200) |

| | | | | | |
|---|---|---|---|---|---|
| grouped | | rt | 4,000 (4,000, 4,000) | 55,700 (51,500, 62,900) | 4,100 (3,700, 4,500) |

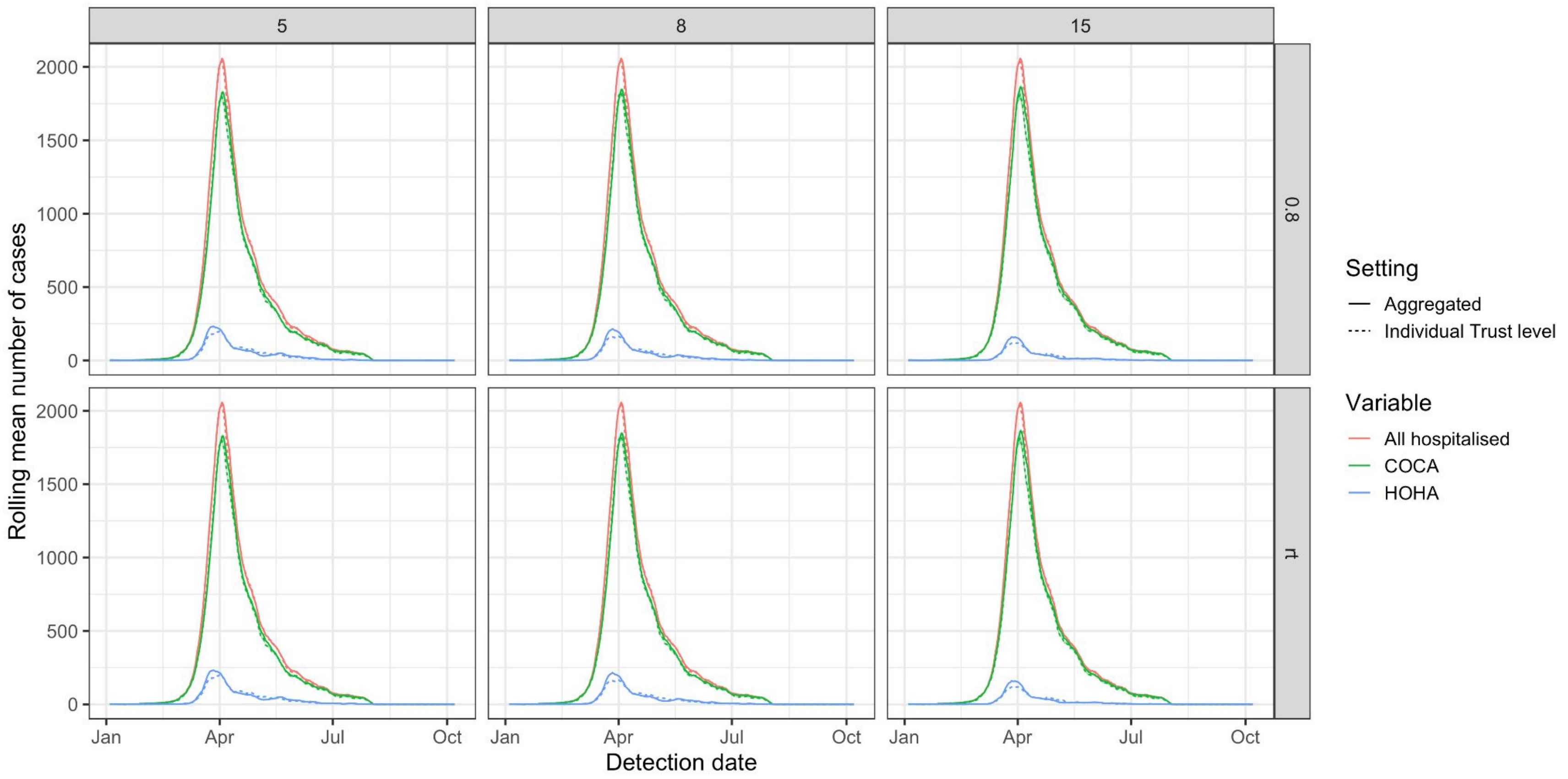


**Figure S15: For the first symptom onset to hospitalisation scenario, there is little variation in the output of key case numbers (all hospitalised (red), community-onset, community-acquired (COCA, green) and hospital-onset, hospital-acquired (HOHA, blue)) if the analysis is performed on the aggregated England setting level (solid line, baseline) or at the individual Trust level and then aggregated (dashed line). The line here is the mean over 200 simulations for the aggregated England setting (50 simulations per Trust for the individual Trust analysis) and 95% range in shaded area.**

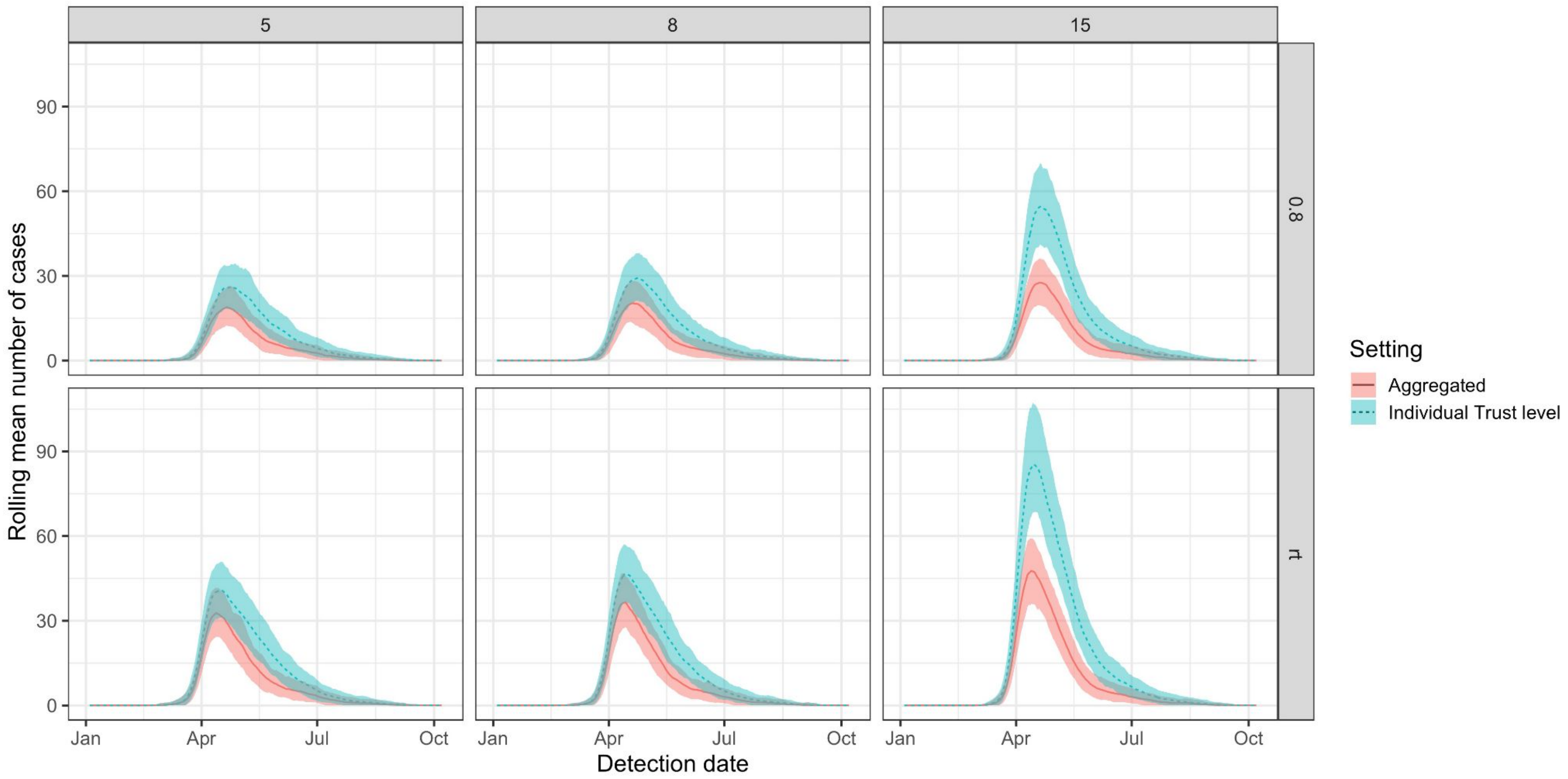


**Figure S16: For the first symptom onset to hospitalisation scenario, there is some variation in the number of community-onset, hospital-linked cases if the analysis is performed on the aggregated England setting level (solid line, baseline, red) or at the individual Trust level and then aggregated (dashed line, blue). The line here is the mean over 200 simulations for the aggregated England setting (50 simulations per Trust for the individual Trust analysis) and 95% range given in the shaded area.**

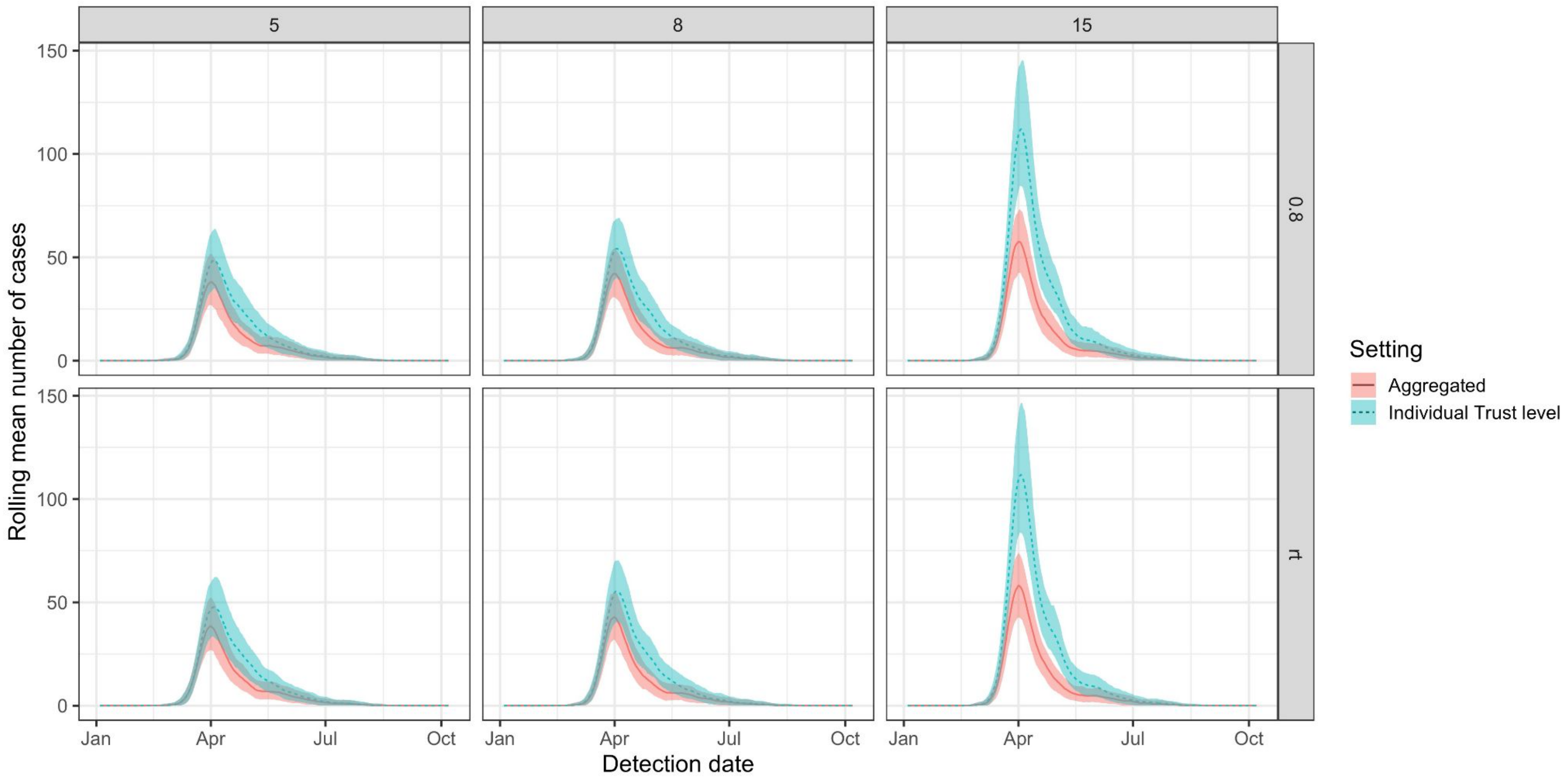


**Figure S17: For the first symptom onset to hospitalisation scenario, there is some variation in the number of community-onset, hospital-acquired cases if the analysis is performed on the aggregated England setting level (solid line, baseline, red) or at the individual Trust level and then aggregated (dashed line, blue). The line here is the mean over 200 simulations for the aggregated England setting (50 simulations per Trust for the individual Trust analysis) and 95% range given in the shaded area.**
